# Microbial diversity modifies the impact of air pollution on pneumococcal disease risk

**DOI:** 10.64898/2025.12.10.25341877

**Authors:** Sophie Belman, Cebile Lekhuleni, Jackie Kleynhans, Giovenale Moirano, Daniela Lührsen, Cristina Carnerero, Susan Meiring, Stephanie W. Lo, Mignon du Plessis, Anne von Gottberg, Rachel Lowe

## Abstract

*Streptococcus pneumoniae* (pneumococcus) remains a major global cause of invasive human disease with seasonal variation suggesting that environmental factors may drive transmission. The pneumococcus includes >100 serotypes and lineages with different disease patterns. Potential interactions between microbial diversity, environmental factors, and invasive pneumococcal disease (IPD) have not been explored. We examined effects of air pollution and meteorological factors on 59,017 IPD cases over 15 years, including 4,350 genome-sequenced isolates, using Bayesian spatio-temporal hierarchical models linking environmental exposures to bacterial diversity. Humidity and air pollution were important environmental drivers. Humidity (33-49%) conferred a 5% disease risk, while high humidity was protective. Weekly PM_2.5_ exposure (50 μg/m³) was associated with a 4% increase in disease risk, was greatest in adults ≥65, and varied by disease type. Quantifying these relationships may refine vaccine-impact models and inform air pollution policy targets.

## Introduction

*Streptococcus pneumoniae* (the pneumococcus) is asymptomatically carried in the human nasopharynx and can cause both non-invasive diseases (such as otitis media and sinusitis), and invasive pneumococcal disease (IPD) such as bacteraemia and meningitis, when the bacterium breaches the epithelium or enters the bloodstream^1,2^. The progression from acquisition, to asymptomatic carriage, to invasive disease is mediated by a combination of environmental drivers, microbial virulence, and host immunity. The pneumococcus has over 800 known genomic lineages, many with hundreds of years of diversity among them. Here we use Global Pneumococcal Sequence Clusters (GPSCs) to classify pneumococcal lineages. The pneumococcus is typically enclosed in a polysaccharide capsule of which there are more than 100 known serotypes. The seven-valent and 13-valent pneumococcal conjugate vaccines (PCV7 and PCV13) target seven and 13 serotypes respectively. This has resulted in serotype replacement by non-vaccine serotypes (NVTs) in the years following vaccine implementation globally^3–6^.

Despite high vaccination coverage, IPD shows consistent seasonal spikes each year^7^. Efforts to reduce residual IPD burden is focused on adapting vaccination strategies to settings with varied sociodemographic contexts and pneumococcal colonization densities^8–11^. However, the role of air quality and meteorological factors, such as air pollution, temperature and humidity in pneumococcal colonization and disease dynamics are typically overlooked. Many geographic regions with the highest pneumococcal disease burden also have populations with extensive exposure to polluted air^12^. Understanding the sensitivity of pneumococcal dynamics to air quality and meteorological factors could allow vaccination strategy modeling and winter hospital capacity preparations to better adapt to different geographies and changing climates. Existing evidence around the impact of environmental factors on pneumococcal disease is heterogenous^13^. Most studies focus on temperature, generally showing that cold temperatures increase disease in temperate climates, although positive associations (higher disease with higher temperatures) have been reported in arid settings^14–16^. Evidence on rainfall and humidity is similarly mixed: while many studies suggest that increasing rainfall and humidity decrease pneumococcal disease incidence, the opposite effect is found in settings such as Senegal^17^, Texas^18^, Spain^16^, and New Zealand^19^ where high levels of rainfall and humidity are associated with increasing disease. Air pollution is widely considered to increase IPD risk, however the mechanisms are poorly understood^20^. A recent scoping review identified only thirteen epidemiological studies (as of January 2026) examining particulate matter (e.g. PM_2.5_ and PM_10_) with just six finding a positive relationship^13,14,19,21–24^. Most relied on simple correlations, few assessed delayed effects (lag times from exposure to outcome), or controlled for confounding meteorological effects^18,19,25^. Overall, important gaps remain, including limited understanding of non-linear and lagged effects, interactions between environmental factors, and variation across age groups, disease presentations, or serotypes. Accounting for underlying pneumococcal diversity when studying the role of environmental factors in driving IPD transmission and incidence is important given the known variability in virulence, carriage durations, and sociodemographic populations affected by different lineages and serotypes^3,7,26^. By accounting for pneumococcal diversity we aim to additionally uncover the biological mechanisms behind environment-microbe interactions.

Here, we first investigate the relationship between a range of air quality and meteorological factors on IPD accounting for delayed and non-linear effects. We then focus on particulate matter <2.5μm in size (PM_2.5_), and the associated risk for IPD while accounting for underlying microbial diversity. We investigate the role of PM_2.5_ in influencing IPD incidence using IPD case data spanning 2005 to 2023 (incorporating genomic data in models through 2019), from 531 hospitals in South Africa. This is a setting with pneumococcal colonization estimates ranging from around 40-60% (typically measured in under 5 year-olds)^27,28^, and marked geographic and climatic diversity. Major public health interventions occurred during the study period (PCV7 introduction in 2009, PCV13 in 2011, and widespread antiretroviral therapy (ART) for treatment of people living with HIV (PLHIV), since 2005)(Table 1). We include underlying pneumococcal phenotypic and genomic data to understand interactions with underlying microbial ecology, accounting for different serotypes and genomic lineages. We apply Bayesian hierarchical models to capture spatial and temporal variability, incorporate delayed and non-linear relationships between environmental exposure and disease onset, and allow for stratification by age group and disease type. These models enable a better understanding of the interaction between environmental factors and pneumococcal populations which play a role in IPD morbidity and mortality.

**Table 1.** Vaccine implementation year, schedule, and serotypes in South Africa. 2+1 indicates a 2+1 schedule whereby children receive the first two doses at 6 and 14 weeks of age and then a booster dose at 9 months.

| **Vaccine**<br>**(introduction year in South Africa and schedule)** | **Serotypes** |
| --- | --- |
| PCV7 (2009; 2+1) | 4, 6B, 9V, 14, 18C, 19F, 23F |
| PCV13 (2011, 2+1 ) | 4, 6B, 9V, 14, 18C, 19F, 23F, 1, 3, 5, 6A, 7F, 19A |

## Results

A total of 59,017 identified IPD cases were included from 531 hospitals in South Africa between 2005-2023. Most IPD cases were found in urban centers where the majority of the South African population resides including 41% of total IPD from Gauteng province, 18% from Western Cape province, as well as 11 and 9% from KwaZulu-Natal and Eastern Cape provinces (which contain the major South African cities of Johannesburg, Cape Town, Durban, and Port Elizabeth respectively)(Figure 1A-B). Disease cases included bacteremia, meningitis, and other atypical IPD types such as pericarditis and empyema. The majority of cases occurred in people aged 15-64 years old (62%), followed by children under 6 years of age (23%). 4.8% of cases were from over-64 year olds. There was a well characterized decrease in serotypes that were targeted by PCVs. We observed NVT persistence (Figure 1C) and consistent seasonality in IPD cases. Cases peaked in colder months across the time period coinciding with the seasonality of PM_2.5_ concentration and inversely correlating with relative humidity (Figure 1D)(Figure S1)(Table S1).

**Figure 1.**
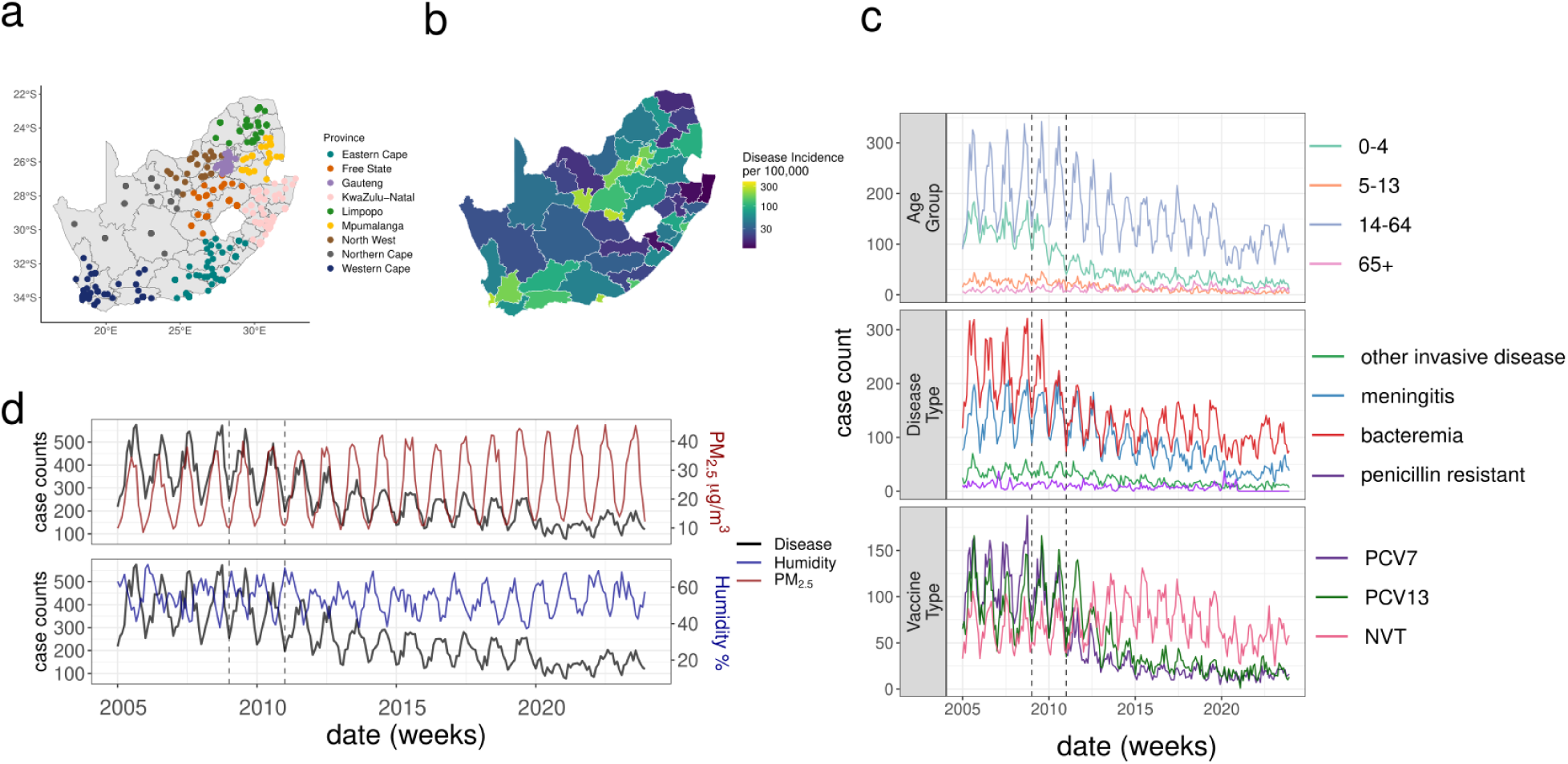
Summary of epidemiological and environmental datasets in South Africa. (a) Hospital locations (n=531) colored by province (n=9) (b) Log10 incidence rate per 100,000 from 2005-2023 by district (n=52) in South Africa (c) monthly case counts by age group, disease type (bacteremia, meningitis, other IPD types, penicillin resistant), and vaccine type (PCV7 = seven-valent vaccine type serotypes; PCV13 = additional six serotypes in the 13-valent vaccine type; NVT = non-vaccine types). (d) monthly pneumococcal disease case counts (black) with mean monthly particulate matter <2.5 μm (PM_2.5_ μg/m^3^) concentration (red), and mean relative humidity (blue) across South Africa from 2005 to 2023; left axis is the monthly case counts, right axis is the concentration (μg/m^3^) of particulate matter <2.5 μm (upper panel) and percent relative humidity (lower panel). Vertical dashed lines in (c) and (d) indicate years of PCV7 implementation (2009) and PCV13 (2011).

### Sociodemographic and Spatiotemporal Effects

We developed a sociodemographic base model which included spatial autocorrelation across districts, seasonal effects, and interannual random effects replicated across provinces, as well as pneumococcal vaccine implementation periods and population density (Table 4). We found similar effects for models spanning 2005-2023 and 2005-2019 but due to the dramatic decrease in IPD cases during the COVID-19 pandemic period (2020 onwards) we proceeded with models restricted to the 2005-2019 period throughout (Table S2)(Table S3). These models were fitted to weekly invasive disease incidence across South Africa district levels (n=52)(Figure S1A-B). The Gauteng and Western Cape provinces had a higher risk of IPD than the national average. The relative risk (RR) derived from the spatial effect for Gauteng province was 13.35 (95% credible interval (CI) 4.19-44.34) and for Western Cape was 1.54 (95% CI 0.84–2.70), representing the area specific risk relative to the average across all provinces. The risk in Western Cape was largely driven by the increased risk in the Cape Town district alone (relative to all district average)(RR: 10.16; 95% CI 7.89-13.09). All districts in Gauteng province exhibited elevated disease risk relative to the national average (RR>1). Across the other provinces, districts which host major cities and transport hubs had twice the risk of IPD compared to average. Some of the exacerbated risk in the main cities, such as those around Johannesburg in Gauteng, may reflect travel to hospitals from more rural areas (Figure 2A)(Figure S1C)(Table S3). There was a seasonal effect with an increased disease risk between the months of May and October with peak risk in August (RR: 1.37; 95% CI 1.33-1.41). In the interannual effect we found a reduction in risk over time even while accounting for vaccination for Gauteng, Free State, Mpumalanga, KwaZulu-Natal, and North West provinces (Figure 2C)(Table S3). This may be attributed to ART uptake also being influential there (Figure S1D).

**Figure 2.**
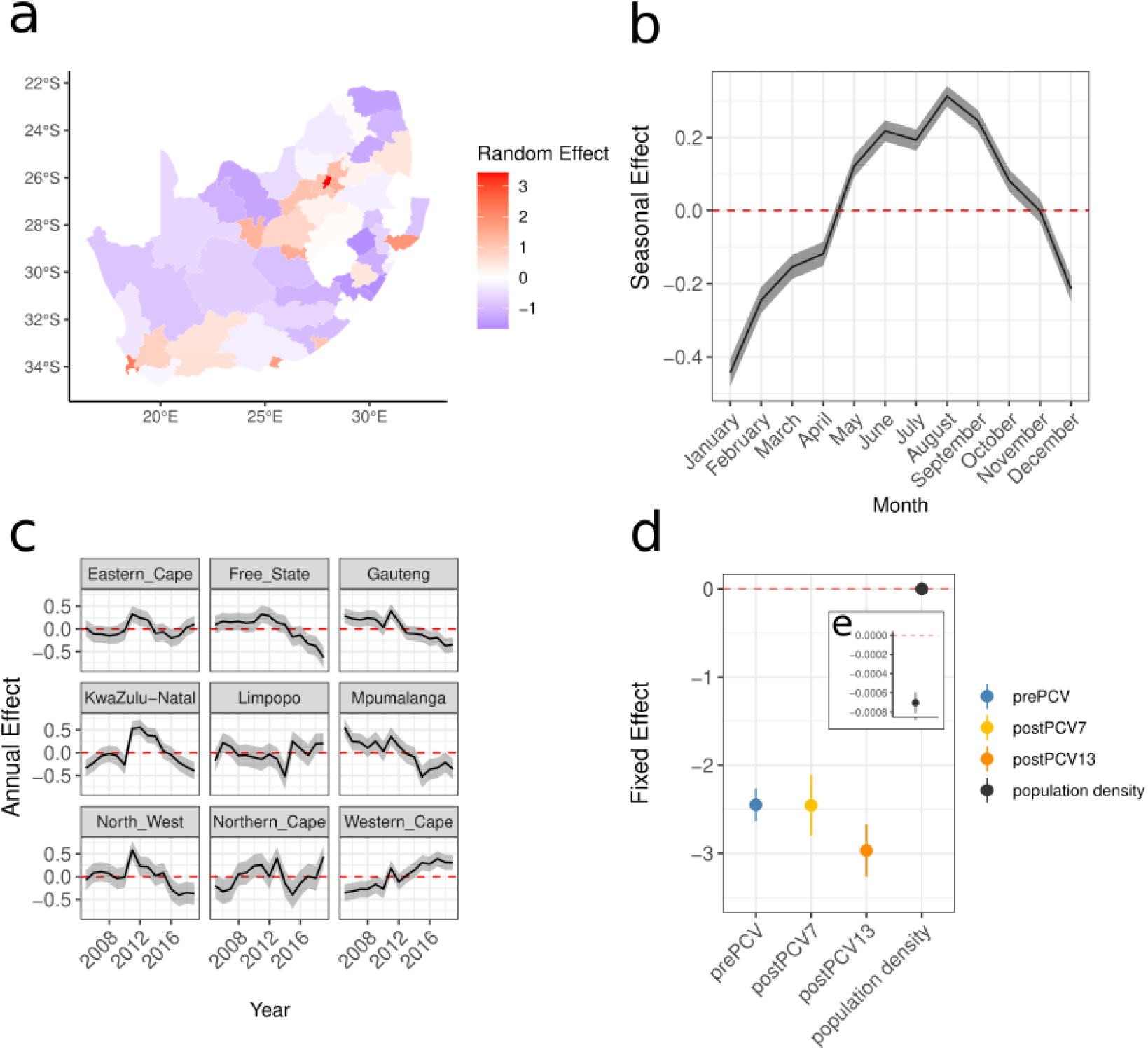
Spatial, seasonal, and interannual effects from the sociodemographic base model. (a) spatial random effects across 52 districts where a positive effect (more cases than expected on average) is red and a negative effect (fewer cases than expected on average) is blue, (b) monthly random effects across the 12 months of the year, the red line denotes effect size of 0, (c) yearly random effects 2005-2019 replicated across the 9 provinces of South Africa (d) median fixed effects of vaccination for each vaccine period pre-PCV7 (2005-2008) (blue), post-PCV7 (2009-2011) (yellow), and post-PCV13 (2012-2019) (orange), and median population density fixed effect (black) (e) the same population density fixed effect shown in d but with y-axis reduced (-8×10^-4^,0) to highlight the effect.. The shading and error bars represent 95% credible intervals.

In developing the sociodemographic base model we examined the impact of ART and vaccination coverage. They were correlated (Pearson *R*^2^ = 0.84) and model formulations which included them had reduced interannual effect variance (Figure S1D)(Figure S2A). We found no effect from ART or PCV on NVT serotypes alone in the 2005-2019 period. A lack of PCV effect on NVTs is expected, as the vaccine does not protect against these serotypes. In contrast, ART is expected to confer protection against all pneumococcal serotypes by improving immune function in PLHIV, particularly among adults, who represent the group most likely to be receiving ART in this high HIV-burden setting^30^. In the 2005-2019 period, NVT IPD cases were skewed toward children and older adults but extending the analysis to 2020-2023 allowed an ART effect on NVTs to emerge. In this time period there were a higher number of IPD cases among adults aged 18-64 years who in this setting are the primary recipients of ART. This increase in adult cases may result from the resurgence of IPD after the COVID-19 pandemic and a gradual decline in indirect vaccine protection over time (Figure S2C-E)^29–33^. Due to the nuanced impact of ART, and the established direct relationship between pneumococcal vaccination and IPD, we included the three PCV periods (pre-PCV7, post-PCV7, and post-PCV13) as the intervention-covariate in the final sociodemographic base model (Equation 1)(Figure 2D)(Table S4).

### Associations between environmental factors and pneumococcal disease risk

We incorporated environmental factors in our sociodemographic base model (described above) to estimate the risk of IPD from environmental exposure across an 8-week time period (Table 4, M12). We included an 8-week lag period due to a previous epidemiological evidence of up to a 1-month lag time from environmental exposure to pneumococcal disease effects, finding the maximum correlation from environmental exposure to disease up to about a 5-week lag time, and previous evidence of a 30 day estimated generation time for the pneumococcus (Figure S3)^6,18,20,34^. Specifically, we used distributed lagged non-linear models (DLNMs) to incorporate the delayed and non-linear effects of environmental factors up to 8-weeks after exposure (Figure S4).

#### Meteorological models

Our meteorological models comprise the sociodemographic base model plus a DLNM for each variable. We included the weekly average of the daily mean, minimum, or maximum temperature. Temperature had a non-linear cumulative association with IPD across the time period with only cold temperatures (average minimum weekly temperatures of 4–10°C) conferring an increased cumulative risk of IPD (6°C cumulative Relative Risk (RR) 1.04: 95% CI 1.01-1.06)(Figure 3A). Warmer temperatures (weeks with an average maximum temperature of 31°C) were associated with a 5% increased risk of IPD relative to the median, rising to an 11% increased RR at 38°C. Similarly, mean weekly temperatures of 24°C were associated with a 5% risk increase (lag week 0 RR 1.05: 95% CI 1.03-1.08)(Figure S5C).

**Figure 3.**
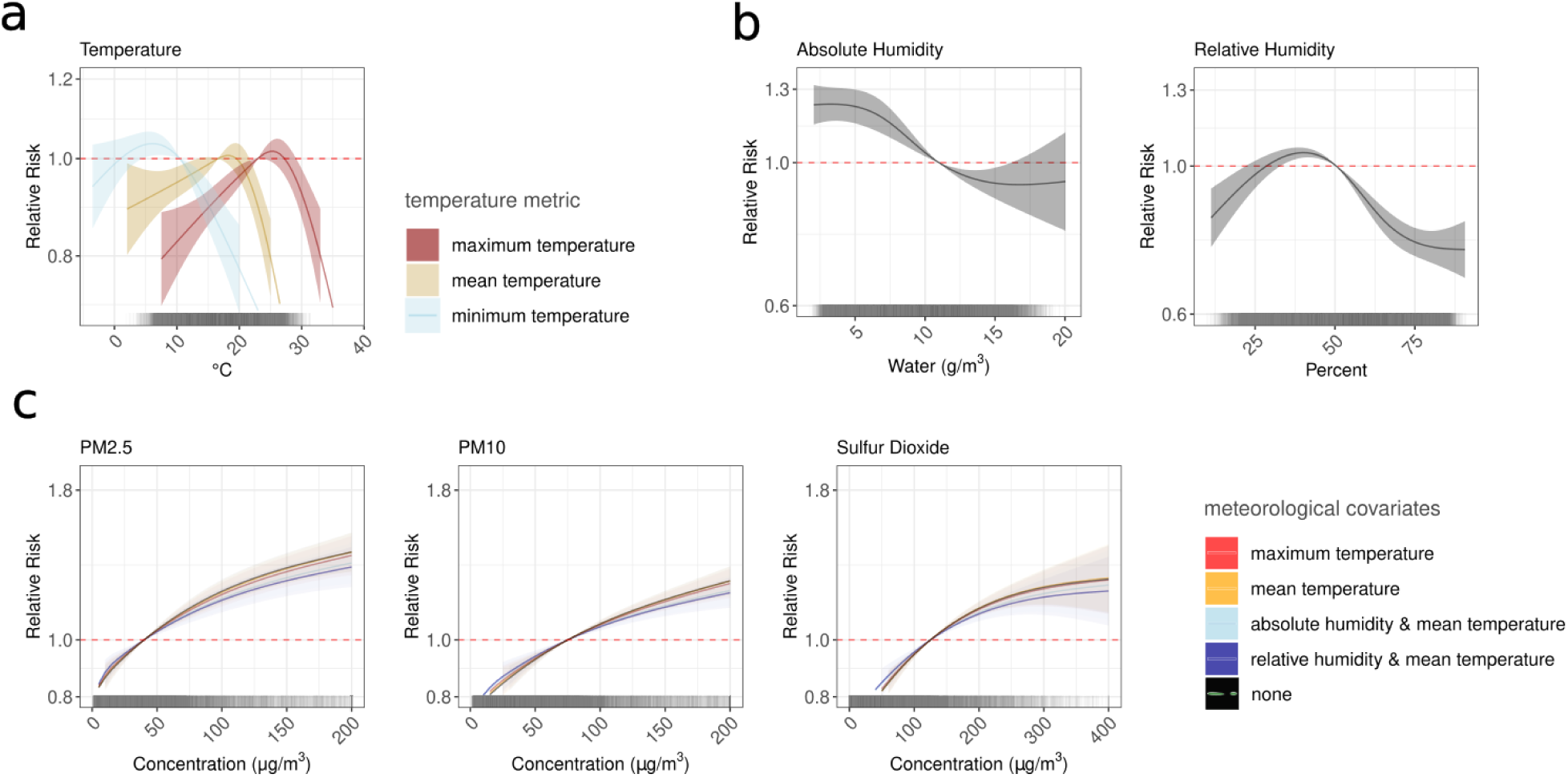
Cumulative relative risk across 8-weeks of lags for (a-b) meteorological models, and (c) air pollution models. (a) Exposure response curves for maximum (pink), mean (yellow), and minimum (blue) temperature in Celsius (℃). (b) Exposure response curves for absolute humidity (g/m^3^) (left) and % relative humidity (right)(c) Exposure response curves for PM_2.5_ (left) PM_10_ (middle) or sulfur dioxide (SO_2_)(right) including models with combinations of meteorological covariates (temperature metrics with linear formulations and humidity metrics with non-linear formulations): maximum temperature (red), mean temperature (yellow), absolute humidity and mean temperature (light blue), relative humidity and mean temperature (dark blue), and none (black) with three week lags for humidity and no lag for temperature. These models are fit to weekly district level data from 2005-2019 across South Africa. The DLNM crossbasis for the humidity variables has 3-degrees of freedom while the rest of the DLNMs have 2-degrees of freedom. At the bottom of (a-d) is a rug plot with the distribution of the variables in the DLNM. For (a) the rug plot is for mean temperature. The shading represents 95% credible intervals.

Relative humidity provided a better model fit (difference in Watanabe–Akaike Information Criterion; ΔWAIC: 77.7) than absolute humidity (the absolute amount of vapor in air g/m^3^) which in contrast to relative humidity does not account for the temperature-dependent variation in the air’s capacity to retain water. Absolute humidity had a stronger association at low values (4 g/m^3^ cumulative RR 1.23: 95% CI 1.16-1.30) with an acute increase in disease risk until a 2-week lag time. For relative humidity the cumulative risk was non-linear. The highest risk occurred at moderate humidity levels (34%- 47%) with a maximum cumulative RR of 1.05 (95% CI 1.03–1.07) at 39.8% relative to the median (Figure 3B). There was a strong protective effect of high relative humidity with cumulative RR decreasing to 0.77 (95% CI 0.73-0.88) at 74.8% relative humidity (Figure 3B). These differences in apparent effect magnitude between absolute and relative humidity should be interpreted cautiously, as they depend on the distribution of each variable. The weaker relative effects observed for relative humidity likely reflect its dependence on temperature, suggesting that temperature partially mediates humidity-related effects (Figure 3B)(Figure S5)(Figure S21)(Table S5).

Increasing precipitation and a higher Standardised Precipitation and Evapotranspiration Index (SPEI) (suggesting reduced drought conditions) decreased IPD risk (negative associations) (Figure S6A-B). Increasing average wind speed increased cumulative IPD risk (positive association), while increasing ozone concentrations (ozone increases with increasing temperatures) decreased cumulative IPD risk; concordant with decreasing risk in the summer months (Figure S6C-D)(Figure S7).

#### Air pollution models

We estimated the risk of disease above the 2016-2029 National Ambient Air Quality Standards (NAAQS) thresholds in South Africa^35^ for PM_2.5_ (40 μg/m^3^), PM_10_ (75 μg/m^3^), and SO_2_ (125 μg/m³)(Table 2)(Table 3). To account for meteorological confounding in air pollution models, we evaluated multiple formulations and lag structures of temperature and humidity. The final air pollution models adjusted for mean temperature in the same week as the pollutant and absolute humidity at lag 3 (three weeks prior)(Table 5)(Table S5)(Figure S8). PM_2.5_, PM_10_, and SO_2_ all had positive associations with IPD (Figure 3C)(Figure S5).

**Table 2.** Description of air quality reanalysis products. Calculations to adjust each air-quality reanalysis product to μg/m^3^ detailing the mass used for PM_2.5_, PM_10_, O_3_, and SO_2_.

| Variable | CAMS (50km<sup>2</sup>) |  | MERRA (80km<sup>2</sup>) |  | MERRA_BiasFix (80km<sup>2</sup>) |  |
| --- | --- | --- | --- | --- | --- | --- |
| | Initial | Adjust<sup>#</sup> to $\mu\text{g}/\text{m}^3$ | Initial | Adjust<sup>#</sup> to $\mu\text{g}/\text{m}^3$ | Initial | Adjust to $\mu\text{g}/\text{m}^3$ |
| PM<sub>2.5</sub> | kg m<sup>-3</sup> | *10<sup>9</sup> | kg m<sup>-3</sup> | *10<sup>9</sup> | kg m<sup>-3</sup> | *10<sup>9</sup> |
| PM<sub>10</sub> | kg m<sup>-3</sup> | *10<sup>9</sup> | kg m<sup>-3</sup> | *10<sup>9</sup> | - | - |
| O<sub>3</sub> | kg kg<sup>-1</sup> | *2.14*10<sup>9</sup> | kg kg<sup>-1</sup> | *2.14*10<sup>9</sup> | - | - |
| SO<sub>2</sub> | kg kg<sup>-1</sup> | *2.63*10<sup>9</sup> | kg kg<sup>-1</sup> | *2.63*10<sup>9</sup> | - | - |
| # air density volume included as 1.23, O<sub>3</sub> density as 2.14 and SO<sub>2</sub> as 2.63 |  |  |  |  |  |  |

**Table 3.** Correlations between observational station data and each air quality analysis product. Relationship between reanalysis products from Copernicus (CAMS), MERRA-2 from NASA, MERRA-2 with a bias adjustment and a machine learning gapless product at 1km grid squares from Wei et al. 2023. We include a spearman’s correlation, mean absolute error (μg/m^3^) and the root mean squared error (μg/m^3^).

|  | Gauteng |  |  |  | KwaZulu-Natal |  |  |  | Gert Sibande, Mpumalanga |  |  |  |
| --- | --- | --- | --- | --- | --- | --- | --- | --- | --- | --- | --- | --- |
| Statistic | CAMS | MERRA-2 | MERRA-2<br>-adjusted | ML<br>Gapless<br>1km | CAMS | MERRA-2 | MERRA-2<br>-adjusted | ML<br>Gapless<br>1km | CAMS | MERRA-2 | MERRA-2<br>-adjusted | ML<br>Gapless<br>1km |
| Correlation | 0.88 | 0.49 | 0.58 | 0.82 | 0.58 | 0.52 | 0.17 | 0.08 | 0.08 | 0.30 | -0.21 | 0.27 |
| MAE | 43.06 | 74.79 | 62.05 | 55.52 | 32.49 | 52.98 | 50.74 | 48.15 | 74.55 | 92.95 | 83.27 | 83.40 |
| RMSE | 58.46 | 81.02 | 67.47 | 61.40 | 42.34 | 59.69 | 57.50 | 55.69 | 105.45 | 119.56 | 113.56 | 111.66 |

**Table 4.** Sociodemographic model formulations with increasing complexity to establish model base at a weekly temporal resolution and including years 2005-2019. WAIC = Watanabe-Akaike Information Criterion, MAE = Mean Absolute Error, CPO = log10 conditional predictive ordinate, and R^2^ likelihood ratio compared to the intercept model. M1-M12 and M16 are in Table S2 and M13-M15 are in Table S4. M12 is the sociodemographic base model used in subsequent environmental model formulations. *f* indicates a non-linear function.

| Model | Formula | WAIC | MAE | CPO | $R^2$ |
| --- | --- | --- | --- | --- | --- |
| M0 | $\log(\rho_{s,t}) = \alpha$ | 99687.81 | 1.18 | 1.23 | 0 |
| M1 | $\log(\rho_{s,t}) = \alpha + \delta_{m(t)} + \gamma_{a(t)}$ | 97301.75 | 1.14 | 1.2 | 0.06 |
| M2 | $\log(\rho_{s,t}) = \alpha + \delta_{m(t)} + u_s + v_s$ | 84082.5 | 0.9 | 1.04 | 0.32 |
| M3 | $\log(\rho_{s,t}) = \alpha + \gamma_{a(t)} + u_s + v_s$ | 82439.36 | 0.8 | 1.02 | 0.35 |
| M4 | $\log(\rho_{s,t}) = \alpha + \delta_{m(t)} + \gamma_{a(t)} + u_s + v_s$ | 80883 | 0.76 | 1 | 0.37 |
| M5 | $\log(\rho_{s,t}) = \alpha + f(pcv_{2009vaccine}) + \delta_{m(t)} + \gamma_{a(t)} + u_s + v_s$ | 80882.95 | 0.76 | 1 | 0.37 |
| M6 | $\log(\rho_{s,t}) = \alpha + f(\text{pcv}_{\text{periods}}) + \delta_{m(t)} + \gamma_{a(t)} + u_s + v_s$ | 80882.89 | 0.76 | 1 | 0.37 |
| M7 | $\log(\rho_{s,t}) = \alpha + f(\text{pcv}_{\text{periods}}) + f(\text{covid}) + \delta_{m(t)} + \gamma_{a(t)} + u_s + v_s$ | 80883.01 | 0.76 | 1 | 0.37 |
| M8 | $\log(\rho_{s,t}) = \alpha + f(\text{pcv}_{\text{periods}}) + \delta_{m(\text{pr}(s),t)} + \gamma_{a(t)} + u_s + v_s$ | 80794.05 | 0.75 | 1 | 0.38 |
| M9 | $\log(\rho_{s,t}) = \alpha + \delta_{m(t)} + \gamma_{\text{pr}(s),a(t)} + u_s + v_s$ | 78943.71 | 0.7 | 0.97 | 0.41 |
| M10 | $\log(\rho_{s,t}) = \alpha + f(\text{pcv}_{2009\text{vaccine}}) + \delta_{m(t)} + \gamma_{\text{pr}(s),a(t)} + u_s + v_s$ | 78942.09 | 0.7 | 0.97 | 0.41 |
| M11 | $\log(\rho_{s,t}) = \alpha + f(\text{pcv}_{\text{periods}}) + \delta_{m(t)} + \gamma_{\text{pr}(s),a(t)} + u_s + v_s$ | 78939.5 | 0.7 | 0.97 | 0.41 |
| **M12** | **$\log(\rho_{s,t}) = \alpha + \beta_{\text{pd}} \text{pd}_{s,a(t)} + f(\text{pcv}_{\text{periods}}) + \delta_{m(t)} + \gamma_{\text{pr}(s),a(t)} + u_s + v_s$** | **78766.03** | **0.7** | **0.97** | **0.41** |
| M13 | $\log(\rho_{s,t}) = \alpha + \beta_{\text{pcv}} \text{pcv}_{\text{coverage},a(t)} + \delta_{m(t)} + \gamma_{\text{pr}(s),a(t)} + u_s + v_s$ | 78940.44 | 0.70 | 0.97 | 0.41 |
| M14 | $\log(\rho_{s,t}) = \alpha + \beta_{\text{art}} \text{art}_{\text{coverage},a(t)} + \delta_{m(t)} + \gamma_{\text{pr}(s),a(t)} + u_s + v_s$ | 78939.15 | 0.70 | 0.97 | 0.41 |
| M15 | $\log(\rho_{s,t}) = \alpha + \beta_{\text{art}} \text{art}_{\text{coverage},\text{pr}(s),a(t)} + \delta_{m(t)} + \gamma_{\text{pr}(s),a(t)} + u_s + v_s$ | 78938.82 | 0.70 | 0.97 | 0.41 |
| M16 | $\log(\rho_{s,t}) = \alpha + \beta_{\text{art}} \text{art}_{\text{coverage},\text{pr}(s),a(t)} + f(\text{pcv}_{\text{periods}}) + \delta_{m(t)} + \gamma_{\text{pr}(s),a(t)} + u_s + v_s$ | 78938.55 | 0.70 | 0.97 | 0.41 |
| M17 | $\log(\rho_{s,t}) = \alpha + \beta_{\text{pd}} \text{pd}_{s,a(t)} + f(\text{pcv}_{\text{periods}}) + \delta_{m(t)} + u_{s(t)} + v_{s(t)}$ | 77310.32 | 0.68 | 0.95 | 0.44 |

📄 Tables\_Env\_SA\_V3

**Table 5.** Model comparison for PM_2.5_ including different model formulations for inclusion of meteorological confounders. These all include the sociodemographic base model and a PM_2.5_ crossbasis (CB_aq_(aq_s,t−ℓ_)) as well as a temperature covariate with no lag time (temp_s,t_) or a 3-week lag time (temp_s,t-3_), and a non-linear formulation of absolute humidity with a 3-week lag time (f_2_(hum_s,t-3_)). Additional evaluated formulations are included in Table S5. *f* indicates a non-linear function.

| Model | Cumulative<br>8-week RR at<br>50μg/m<sup>3</sup> | WAIC | MAE | CPO |
| --- | --- | --- | --- | --- |
| $\log(\rho_{s,t}) = \alpha + \beta_{pd}pd_{s,a(t)} + f(pcv_{periods}) + CB_{aq}(aq_{s,t-l}) + \delta_{m(t)} + \gamma_{pr(s),a(t)} + u_s + v_s$ | 1.04 (1.03-1.05) | 78684 | 0.697 | 0.970 |
| $\log(\rho_{s,t}) = \alpha + \beta_{pd}pd_{s,a(t)} + f(pcv_{periods}) + \beta_{temp}temp_{s,t-3} + CB_{aq}(aq_{s,t-l}) + \delta_{m(t)} + \gamma_{pr(s),a(t)} + u_s + v_s$ | 1.04 (1.03-1.05) | 78686 | 0.697 | 0.970 |
| $\log(\rho_{s,t}) = \alpha + \beta_{pd}pd_{s,a(t)} + f(pcv_{periods}) + f(hum_{s,t-3}) + CB_{aq}(aq_{s,t-l}) + CB_{aq}(aq_{s,t-l}) \cdot temp_{s,t-3} + \delta_{m(t)} + \gamma_{pr(s),a(t)} + u_s + v_s +$ | 1.04 (1.01-1.007) | 78662 | 0.696 | 0.970 |
| $\log(\rho_{s,t}) = \alpha + \beta_{pd}pd_{s,a(t)} + f(pcv_{periods}) + \beta_{temp}temp_{s,t-3} + f(hum_{s,t-3}) + CB_{aq}(aq_{s,t-l}) + \delta_{m(t)} + \gamma_{pr(s),a(t)} + u_s + v_s$ | 1.03 (1.02-1.05) | 78661 | 0.696 | 0.970 |
| $\log(\rho_{s,t}) = \alpha + \beta_{pd}pd_{s,a(t)} + f(pcv_{periods}) + f(hum_{s,t-3}) + CB_{aq}(aq_{s,t-\ell}) + CB_{temp}(temp_{s,t-\ell}) + \delta_{m(t)} + \gamma_{pr(s),a(t)} + u_s + v_s$ | 1.03 (1.02 - 1.05) | 78659 | 0.696 | 0.970 |
| $\log(\rho_{s,t}) = \alpha + \beta_{pd}pd_{s,a(t)} + f(pcv_{periods}) + f(hum_{s,t-3}) + CB_{aq}(aq_{s,t-\ell}) + CB_{aq}(aq_{s,t-\ell}) \cdot temp_{s,t} + \delta_{m(t)} + \gamma_{pr(s),a(t)} + u_s + v_s$ | 1.03 (1.02-1.05) | 78658 | 0.696 | 0.970 |

With a weekly average of 50 µg/m³ of PM_2.5_ there was a 3.4% 8-week cumulative increased risk of IPD (cumulative RR 1.03: 95% CI 1.02-1.05). Strikingly, there was a 18% increased IPD risk (cumulative RR 1.18: 95% CI 1.12-1.23) with a weekly PM_2.5_ of 100 µg/m³ (Figure 3C) The risks from high (100 µg/m³) concentrations of PM_2.5_ were immediate (lag week 0 RR: 1.02 (95% CI 1.01-1.04) and persisted for 6 weeks (Figure S5). These patterns were robust to models fitted to Gauteng province alone, and nationally, excluding Gauteng (Figure 3A)(Figure S9-S12)(Table S5). We also explored the potential for fine scale spatiotemporal confounding of the PM_2.5_ effect by allowing spatial effects to vary by year. This showed high correlation with the interannual province level effects from the sociodemographic base model, with a few urban districts as outliers. However, the PM_2.5_ exposure-response relationship was robust to this analysis indicating that fine-scale spatiotemporal confounding was not influential (Figure S13). The City of Johannesburg regularly exceeded weekly averages of 100 µg/m³ of PM_2.5_ (32 weeks in 2023) and across South Africa the weekly PM_2.5_ concentration was at least 50 µg/m³ in about 23 districts a year (about 14 weeks per district)(Figure S14)(Table S5). In 2019 Gauteng province had a mean PM_2.5_ concentration of 117 µg/m³ and 718 observed IPD cases, and the population attributable fraction, based on weekly PM_2.5_ concentrations was 2.19%. This implies that if PM_2.5_ had been maintained at or below 40 µg/m³ 16 of the 718 cases could theoretically have been averted; if this were reduced further, to the more stringent WHO daily threshold of 15 µg/m³, 24.8 cases (3.45%) would be averted.

A weekly PM_10_ concentration of 100 μg/m³ increased the cumulative 8-week risk of disease 5.6% (cumulative RR 1.06: 95% CI 1.04-1.07) and had a lower acute risk (lag week 0) than for PM_2.5_ (Figure S5). With a weekly SO_2_ concentration of 150 μg/m³ (target threshold in South Africa 125 µg/m³) the risk of IPD only began 3 weeks after exposure and remained persistent up to 8 weeks after exposure (weekly SO_2_ of 180 μg/m³ cumulative RR 1.09: 95% CI 1.06-1.11)(Figure S5)(Figure S12).

### Risk from PM_2.5_ varies by age, disease, and pneumococcal serotype

To assess the varying impact of air pollution across outcome types we stratified IPD outcomes by particular demographic groups, disease outcomes, and pneumococcal serotypes. We employed the same air pollution model formulation described above which included the sociodemographic base model with meteorological confounders (temperature and humidity), and a DLNM for PM_2.5_, PM_10_, and SO_2_ respectively. We focus here on PM_2.5_ due to its majority anthropogenic sourcing in South Africa^36–38^ and clear positive association with pneumococcal disease (Table S5).

#### Age and disease outcome with PM_2.5_

The highest risk from PM_2.5_ was for those aged ≥65 years, with a cumulative RR for 50 μg/m^3^ weekly average PM_2.5_ of 1.16 (95% CI 1.10-1.22)(Figure 4B). The relative risk ratio (RR ratio) at 50 μg/m^3^ PM_2.5_ for ≥65 year olds and all IPD was 1.12 (95% CI 1.07-1.17) indicating a 12% higher risk for this age group at this PM_2.5_ concentration (Figure 4A). PM_2.5_ had a more acute impact on those between the ages of 15-64 with a RR>1 in the same week as high PM_2.5_ concentrations (lag 0) while all other demographic groups showed similar lag patterns with the highest risk 3-4 weeks after exposure. This may reflect the higher prevalence of HIV among 15-64 year olds^33^. The risk for children ≤5 years old was not negligible. For children ≤5 years, at 50 μg/m^3^ weekly average PM_2.5_ there was an 8-week cumulative RR of 1.03 (95% CI 1.01-1.05), and for females of all ages it was 1.03 (95% CI 1.01-1.04). While the risk for children ≤5 years old is similar to the average risk of IPD across the population it translates to 3% more children with IPD at high pollution levels and highlights that PM_2.5_ does not only pose an increased disease risk for adults (Figure S15). Models for particular disease outcomes (bacteremia, meningitis, other invasive disease, and penicillin resistant disease) had similar risk patterns with exposure to weekly average 50 μg/m^3^ PM_2.5_. However, there was a higher cumulative risk (RR 1.12: 95% CI 1.08-1.16) and a more acute risk (>1 at early lag weeks) for other invasive disease types (such as pericarditis or empyema) (Figure 4C)(Figure S15). The risk with a 50 μg/m^3^ weekly average was higher for meningitis (RR 1.05: 95% CI 1.03-1.06) than for bacteremia (RR 1.01: 95% CI 1.00-1.03)(Figure 4C), possibly indicating different mechanisms of effect across IPD types. The risk for IPD with penicillin resistance with 50 μg/m^3^ weekly average PM_2.5_ was 8.9% (RR 1.09: 95% CI 1.03-1.15)(Figure 4C) and a 5% higher risk (RR ratio 1.05: 95% CI 1.00-1.1) for penicillin resistant disease with high PM_2.5_ as compared to the main model (all IPD)(Figure 4A).

**Figure 4.**
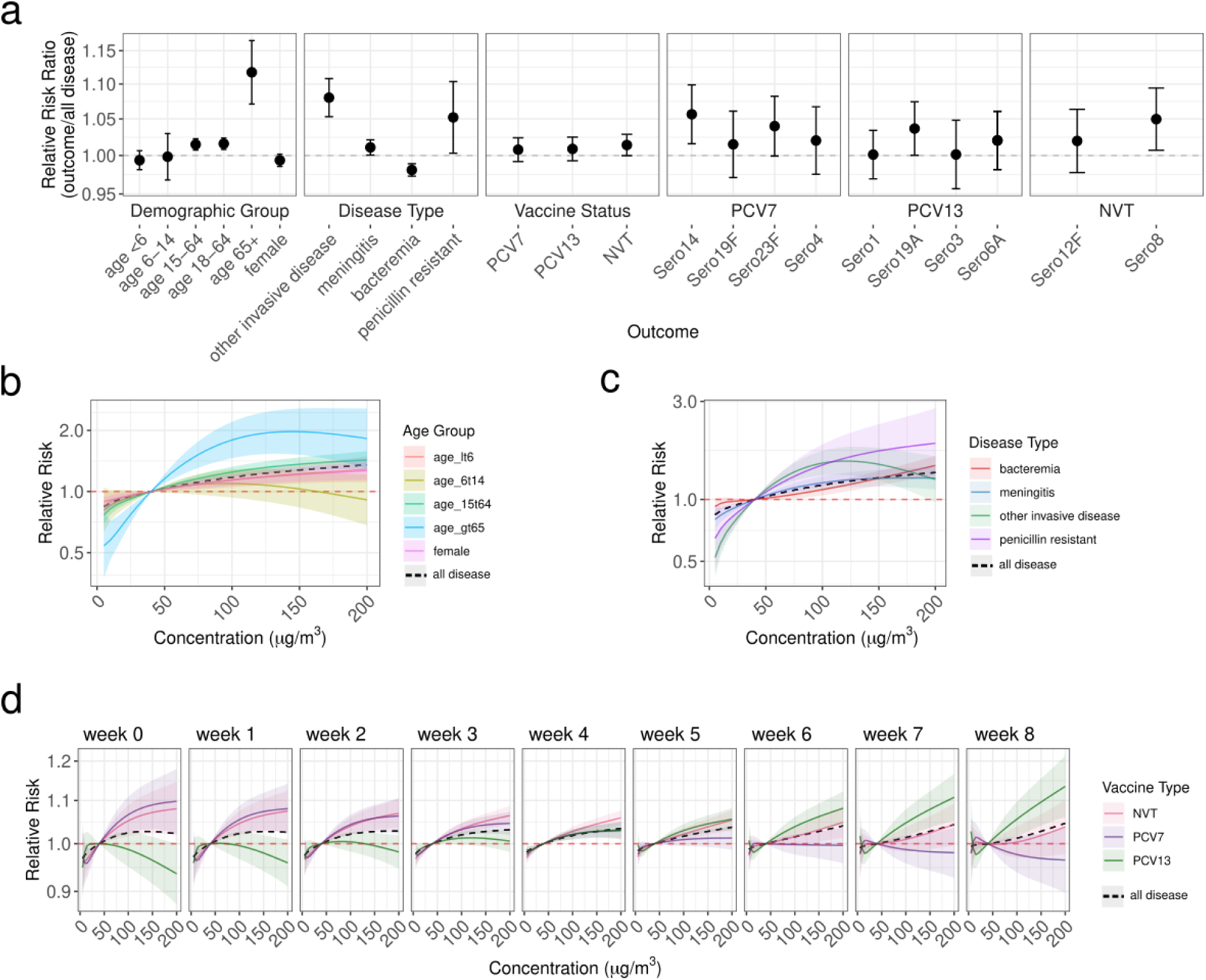
Risk of specific sub-outcomes of invasive pneumococcal disease given weekly averages of particulate matter <2.5μm (PM_2.5_) in the air pollution model framework. These models are all fitted to case data at the district level from 2005-2019. (a) reporting the relative risk ratio of each specific model compared to the model including all disease at 50 μg/m^3^ (exceeding the NAAQS threshold for PM_2.5_ by 10 μg/m^3^) (b) across age groups including age ≤5 (red), age 6-14 (light green), age 15-64 (green), and age 65 and older (blue), and including only females (purple), (c) the relative risk of PM_2.5_ across disease types for bacteremia (red), meningitis (blue), and other invasive disease types (green), penicillin resistant infections (purple) and all disease (black). Weekly lag times for b and c are available in Figure S15. (d) models for disease caused by serotypes targeted by PCV7 (purple), additional 6 serotypes in PCV13 (green), and NVT (red) with a 0 to 8 week lag time from PM_2.5_ to disease for each vaccine type group (pcv7=purple, pcv13=green, nvt=pink) Cumulative vaccine type exposure-response curves are in Figure S16. All models also include the RR from a model including all disease cases (black dashed). The shading and error bars represent 95% credible intervals for (a-d).

#### Serotype and vaccine type with PM_2.5_

The cumulative risk from PM_2.5_ across all vaccine types was similar across the 8 weeks, there was a varied lag pattern with an immediate effect for NVTs and PCV7 serotypes but a longer time to effect for the additional serotypes in PCV13 (Figure 4D)(Figure S16B). These effects persisted when we stratified by pre- and post-PCV periods implying a difference in mechanism beyond vaccine associated changes in serotype proportions (Figure S16A). When further stratified by specific serotypes variable underlying dynamics emerged. PCV7 serotypes 14 and 23F, NVT serotype 8, and PCV13 serotype 19A had RR ratios of >1 with exposure to 50 μg/m^3^ of PM_2.5_ weekly when compared to the main model (Figure 4A)(Figure S17A-B). Notably, and in contrast to other serotypes we found acute (no lag time from exposure) risks from PM_2.5_ for NVT serotype 8 (94% GPSC3 in this dataset), and PCV13 serotypes 23F (68% GPSC14) and serotype 4 (58% GPSC70). These are all known invasive types (Figure S17). Furthermore, the 8-week cumulative risk for serotype 14 disease with 50 μg/m^3^ average weekly PM_2.5_ exposure was greater than on average in the population (RR ratio 1.06: 95% CI 1.02-1.09)(Figure 4A). Serotype 14 is targeted by PCV13 and in South Africa predominantly is represented across several different lineages including the lineages GPSC10 (49%), GPSC18 (25%), and GPSC9 (20%)(Figure S17C). The predominantly lagged effect of PM_2.5_ on serotypes included in the vaccine indicates a protective effect against some of the immediate risks conferred from high levels of PM_2.5_ for IPD (Figure 4D).

### Timing of air quality effects is modified by lineage diversity

We estimated the impact of PM_2.5_ on IPD at the province level according to particular GPSC-lineage prevalence, accounting for a 8-week lag time for the years 2005-2019. In these models, we included IPD as the outcome and incorporated genomic lineages into the model (n=4350) by including the proportion sequenced at each space-time increment as an effect modifier (Figure S18). We interacted the crossbasis for each environmental factor with the proportion of each dominant genomic lineage (n>=100 as well as GPSC8) among sequenced isolates (Figure S19)(Table S1). Models with GPSC41 and GPSC8 had worse goodness-of-fit metrics than the model which excluded lineage diversity interaction effects (Figure S20)(Table S5).

We explored the risk of IPD given the GPSC prevalence across concentrations of PM_2.5_ (where prevalence is across quantiles of GPSC proportions over the entire time period; low=0.1 and high=0.9)(Figure 5A)(Table S6). While the 8-week cumulative effect did not vary with lineage diversity there was variation in the risk across lags and GPSCs (Figure S20A-B)(Figure S22). There was an acute effect for GPSC21 whereby the risk of IPD was elevated given high prevalence of these lineages. The cumulative effect with a GPSC21 interaction also had different exposure-response curves between high and low prevalence (Figure 5C). We found an immediate effect such that locations with a high prevalence of GPSC21 had an acute risk of IPD in the same week as high concentrations of PM_2.5_, whereas times and locations with low prevalence of GPSC21 have an increased risk starting at 5 weeks after the exposure (Figure 5C). In this dataset GPSC21 comprises serotype 19F (n=90) with one instance of 19A in 2005. Serotype 19F is targeted by PCV7 and is characterized in South Africa by an initial GPSC1-lineage (n=114) which decreased post-PCV when GPSC21-19F became predominant (Figure 5B)^7^. When we re-run the GPSC21 interaction models splitting by pre- and post-PCV period we see the effect persisting in the post-PCV period with poor model performance due to low-n in the pre-PCV period (Figure S21). While further investigation is required to understand the mechanism by which GPSC21-19F is causing invasive disease immediately following (same week up to 2 weeks after) high levels of PM_2.5_, it is clear that there are some differences in environmental effects given the circulating lineage diversity.

**Figure 5.**
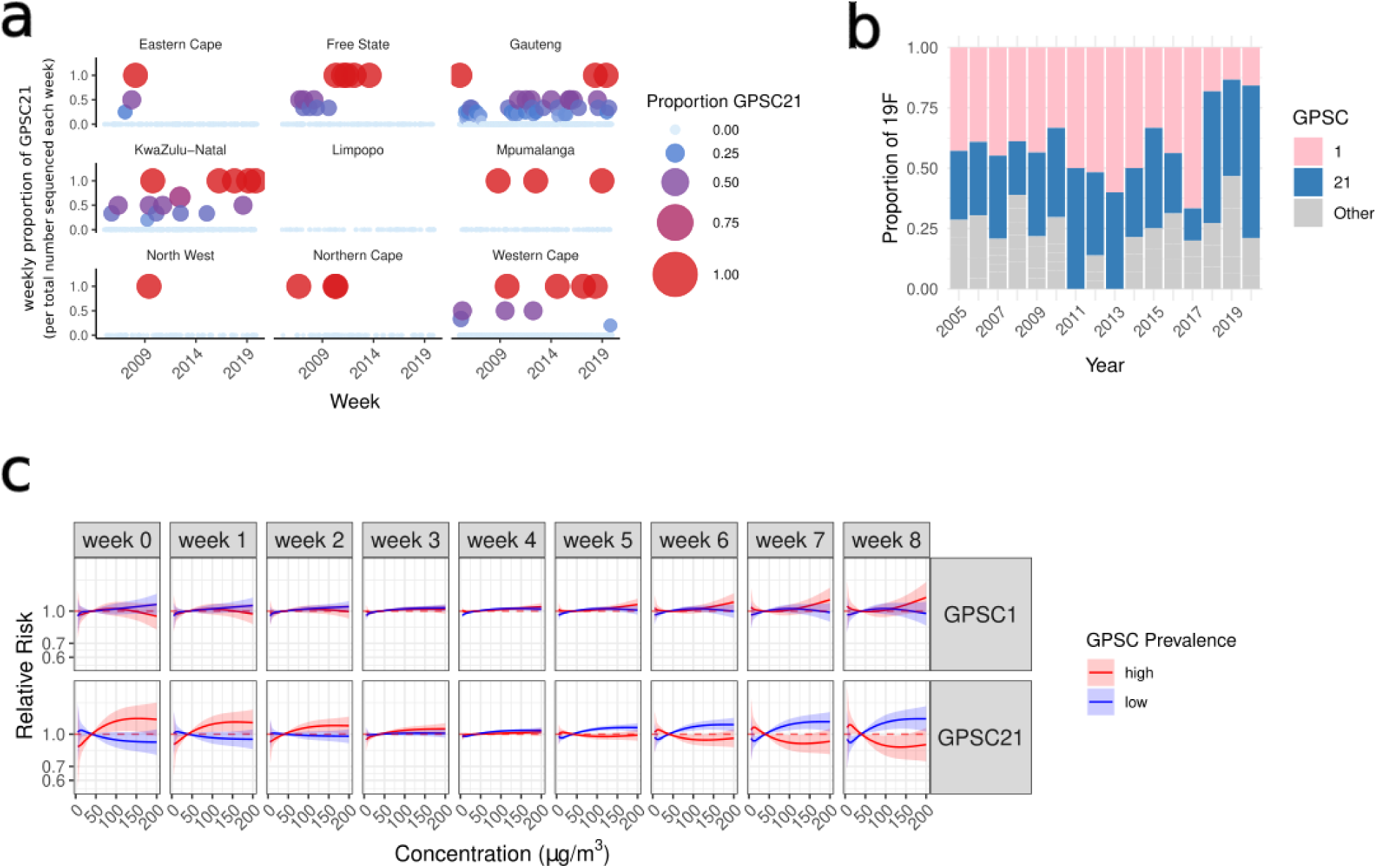
Incorporating GPSC21 by modifying the effect of PM_2.5_ on IPD in the air pollution model framework by (a) the proportion of sequences which were GPSC21 in each week and province per the total number sequenced in each week and (b) the proportion of serotype 19F isolates which are found on each GPSC per year highlighting GPSC1 (pink) and GPSC21 (blue) (c) exposure-response curves for the relative risk of province-level IPD across concentrations of PM_2.5_ for GPSC1 (top) and GPSC21 (bottom) including the curves for high prevalence (red) and low prevalence (blue) of each lineage. The shading represents 95% credible intervals.

## Discussion

In this study, we applied spatio-temporal Bayesian hierarchical models to investigate the relationship between meteorological factors and air pollutant exposure and the risk of IPD, while accounting for microbial diversity and key interventions (ART and PCV). We then focused on the effect of exposure to PM_2.5_ across demographic groups, disease outcomes, serotypes, and lineages. In all presented models we accounted for interannual variability (replicated across provinces), seasonality, spatial autocorrelation, vaccination periods, and population density. We elucidated the effect of meteorological factors and air pollutants on IPD. We focused on PM*_2.5_*, finding variable lag-times and effects across pneumococcal serotypes, age groups, and disease types, and found the greatest increased risk in older adults. We examined whether genomic lineages modified the effects of PM_2.5_ on IPD and found a higher short-term, acute (shortly after exposure) effect of PM_2.5_ when GPSC21-19F was highly prevalent (RR = 1.04; 95% CI 1.01–1.06).

We identified significant protective effects of ART and PCV on IPD although ARTs exacerbated impact on post-2020 NVTs was unexpected. It may reflect the increase in adult IPD cases in that period, and a waning effect of PCV (the major decreases in VT serotypes occurred at earlier time points^7^). Overall the impact of ART on opportunistic infections cannot be understated^33,42–44^. Our study corroborates some previous findings regarding the effects of meteorological factors and IPD, such as increased risk at lower values for humidity, temperature and precipitation, contributing to a growing pool of heterogeneous evidence across different sociodemographic and climatic regions^13^. We find that the protective effect of high humidity is the strongest cumulative 8-week meteorological effect size we identified, providing around a 25% risk reduction for IPD. Effect sizes for temperature variables and air pollution are more modest (below 10%). We focused the study on PM_2.5_ due to sources often being anthropogenic and amenable to action^45^. Our study confirmed a positive relationship between PM_2.5_ and IPD with similar positive associations with PM_10_, and SO_2_. The impact of underlying pneumococcal population diversity on the effect of air pollution has not previously been explored^46^. Notably, when we stratified the models by serotype we found an immediate effect from PM_2.5_ on some specific serotypes including NVT serotype 8 and vaccine serotypes 4 and 23F but found a lagged effect for other serotypes. Different pneumococcal serotypes are known to vary in their invasive potential implying variability in disease mechanism^47^. The mechanism behind the effect of PM_2.5_ on IPD is likely both on the host side (e.g. increased susceptibility to colonization^48^, decreased mucociliary clearance^49^) and the bacterial side (e.g. increased transformation capacity^50^ and biofilm formation^51^)^20^. The immediate risk from PM_2.5_ on NVTs and specific VTs may imply that there is a fitness advantage for these serotypes in high pollution areas. The lagged effect among most VTs suggests that vaccines provide partial protection against immediate PM_2.5_-associated risk. The PM_2.5_ effects persist across all age groups independently. The similarly acute risk for PCV7 and NVT types, compared to the lagged risk for PCV13 types, may reflect different colonization burdens in the population. For example, there is previous evidence that PCV13 serotypes had a larger decreased fitness after vaccination than PCV7 types and that there were greater reductions in PCV13 serotype carriage than for PCV7 serotypes^36^. A possible mechanism for the lagged response to PM_2.5_ among PCV13 types may be that they have lower colonization rates in the population leading to a delayed IPD risk from PM_2.5_^37^. Serotypes with a high burden in populations exposed to elevated air pollution may exhibit more immediate effects, whereas serotypes that are not yet widely colonising may show delayed responses. Evidence on the impact of air pollution on carriage acquisition remains mixed. Several previous studies have identified increased epithelial adhesion, biofilm formation, and bacterial burden with air pollution^46–49^, while others find a decrease in carriage acquisition with increasing PM_2.5_ concentrations^14,51^. The acute effects which we found for serotype 4, 23F, and 19A indicate that there may be a different mechanism for their invasion than other serotypes. Serotype 4 has been particularly notable in causing invasive disease among adults despite its inclusion in the vaccine across the globe^7,52,53^. This may reflect its exacerbation by environmental factors. This serotype has high proportions of the gene *psrP*, an adhesin and a known virulence factor which together with microabrasions from exposure to PM_2.5_^23,24,54^ may increase invasion with no lag time^53^, although this alone is not sufficient given that *psrP* is present across a range of lineages.

The most acute effect (immediate risk from PM_2.5_) is among those aged 15-64 years and the cumulative effect is greatest for those ≥65 years-old. Importantly, there is a 2.7% increased cumulative risk of IPD for children ≤5 years-old with exposure to 50 μg/m^3^ and a 14% increased risk with exposure to 100 μg/m^3^. While this is lower than the risk for adults it is non-negligible. These associations have been previously described for adults and children, with highest risk for older adults^21,23,24^. The increased risk from PM_2.5_ for older adults may reflect a cumulative exposure to air pollution which ultimately results in decreased lung function^53^. This cumulative host-level risk combined with acute exposure events to high levels of PM_2.5_ may trigger increased risk for IPD from PM_2.5_. We did not find any differences when stratifying by sex. However, gendered effects may be masked by the lack of household air pollution in this study^54^.

The pneumococcus is highly diverse and while the serotype data can summarize some of that diversity the immense recombination and dynamic population structure of the pneumococcus cannot be ignored. GPSCs have different carriage durations and demographic propensities^3,4^. It was not possible to directly stratify models including different GPSCs as an outcome, due to the high diversity (low-per-GPSC n), so we instead evaluated the interaction between environmental factors and GPSC prevalence. This allowed us to develop converging models adjusting for the underlying genomic population diversity.

GPSC21-19F is an emergent strain post-PCV in South Africa. The persistent serotype 19F (including PCV) is more frequently expressed by GPSC21 than the pre-PCV GPSC1-19F strain. Serotype 19F is especially prevalent among 5-18 year olds and behavioral differences may contribute to the acute effects of PM_2.5_ on GPSC21-IPD^7^. Direct experimental evidence linking lineage-specific mechanistic data on PM_2.5_-lineage data is currently lacking. Expanding genomic surveillance can support more mechanistic studies to investigate the specific biological interactions which may promote a PM_2.5_ fitness advantage for particular lineages.

The differences in PM_2.5_ effect across serotypes, GPSCs, age groups, and disease outcomes is biologically plausible given the host-pathogen-environment interplay driving disease as well as the massive variability in virulence, carriage duration and burden, genomic content, age group affected, and disease presentation across the different pneumococcal serotypes and lineages^3,5,26^. To further understand the mechanisms by which different serotypes or lineages are affected by PM_2.5_ more experimental evidence, or lineage/serotype stratified epidemiological analysis is necessary. The concentrations of PM_2.5_ and PM_10_ found in South Africa, especially the province of Gauteng, are extremely high. The strong positive association observed in this study is likely partially due to the high exposure concentration. The current daily threshold aim for PM_2.5_ in South Africa is 40 μg/m^3^ whereas the WHO threshold is 15 μg/m^3^. We found in the year 2019 that 15.7 cases might have been averted if the South African threshold was met (24.8 cases by meeting the WHO threshold). More stringent air pollution goals could directly mitigate IPD.

There is an inextricable, bidirectional link between PM_2.5_ and climate change in that particulate matter influences the Earth’s radiative balance and atmospheric chemistry, while changes in temperature, precipitation, and land use affect the formation, composition, and distribution of PM^45,55^. Furthermore, burning of fossil fuels both increases air pollution and global temperatures. There is biological plausibility for air pollution directly impacting host susceptibility, and promoting a fitness advantage for particular microbial traits. Our findings show that high levels of air pollution create conditions that favor particular serotypes and lineages.

Here we provide the first microbial diversity-aware estimates of the risk of IPD driven by air pollution, demonstrating that the risk is not uniform across human or microbial populations with variable effects on at-risk groups and disease types. These interactions should be studied further across different regions with varied climatic conditions and demographic structures. Mechanistic experiments could enhance our understanding of the particular biology leading to these interactions between microbial diversity, host susceptibility, and environment. The time period, high spatial resolution, and extensive metadata of our study is an asset for robust space-time ecological modeling studies. We believe that this novel approach for including genomic data could be leveraged for other infectious diseases to incorporate microbial genomic data into environmental epidemiological models.

### Limitations

The reanalysis and observational datasets for PM_2.5_ are imperfect. As we demonstrate in our comparison, air quality reanalysis models do not correlate well with the observational data across all sites. The available observational data from South African Air Quality Information Services (SAAQIS) across this time period is sparse and limits our ability to select an ideal reanalysis product. The correlation between CAMS and the observations was better than MERRA-2 and other bias corrected datasets. The absence of reanalysis models which are bias corrected to this region, and lack of observational data points for many less-urban regions, highlights the limitations of addressing the environmental impact of high-burden infectious diseases in low-income settings, with uncorrected environmental data^56,57^. To improve understanding of the environment’s impact on high mortality infectious diseases efforts must be made to improve air pollution global reanalysis models^58^. The air quality standards we used as baseline values are the 24-hour standards set by NAAQS. These standards are higher than the health standards set by the WHO due to the high concentration of pollutants in some South African cities. As such, our relative risk estimates are calculated against threshold defined for the South African context but would be higher if compared to the WHO standards. We used weekly means rather than the 24-hour means for air quality metrics so these thresholds are lenient (e.g. if 1 day within a 7 day period has a 24 hour concentration exceeding the threshold but the other 6 days do not, the weekly mean concentration will likely be below the threshold). A limitation of this study is the lack of granular data on indoor air pollution exposure. However, given that indoor and ambient pollutants are often closely correlated, our results likely capture the core exposure-outcome association, though excludes the precise contribution of indoor air pollution^59,60^. We also lacked primary data on indoor household factors, but include population density and spatiotemporal random effects as proxies for some of this unobserved risk. Our inability to account for sub-district level variations in household factors is a further limitation, especially given the known differences in household air pollution exposure when stratified by sex^61^. It is also important to note that extreme rainfall events and flooding may also influence IPD dynamics through mechanisms such as population displacement, water contamination, and disruption of health services^62^. These complex effects were not specifically examined in the present analysis and warrant further investigation.

Environmental reanalysis datasets should be expanded and refined across a wider range of geographies. This may require targeted investment in sustainable air quality monitoring networks, but the resulting evidence would elucidate air quality evolution and the impact on human health globally. Despite our inclusion of the entire GERMS-SA dataset across the time periods, sources of ascertainment bias include access to care, and correct processing and acquisition of specimens. Our data is from hospital location so that in cases of long-distance travel to hospitals cases may be attributed to a spatial grid different from the patient’s home location. While audits were conducted to capture unreported cases these excluded serotype data, and despite our study including a large proportion of genomic sequences these did not encompass the entire epidemiological dataset. The isolates selected for genomic sequencing were randomly selected according to specific age groups in each sequencing set (as described in the methods). By incorporating the genomic lineages as a modifying proportional interaction effect, and normalizing this by the total number of sequences in each space-time increment we try to limit these biases. We did not include carriage isolates in this analysis, as the focus of GERMS-SA is on invasive disease. Consequently, our results pertain only to invasive pneumococcal disease (IPD) lineages and serotypes. This also means that cases of non-bacteraemic pneumonia are not captured in this study. This exclusion may be skewing the impact of some of our environmental associations as a whole. Understanding the relationships between environmental factors and bacteremic versus non-bacteremic pneumonia is an important direction for future work.

### Conclusion

In summary, environmental factors influence IPD risk in South Africa with temperature, humidity, and air pollution posing the highest risk. The relationships we found are delayed and non-linear, and vary given the serotypes or lineage circulating. While there was a strong association between PM_2.5_ and IPD in all age groups independently, the highest risk is among those ≥65, and the most immediate risk is for those between 15 and 64. The lagged and variable effects indicate that the mechanism by which PM_2.5_ increases IPD risk varies across groups (e.g., ages, serotypes, immunity). This study provides strong evidence that diverse bacterial populations are not uniformly affected by the environment. IPD patterns are influenced by increasing urban populations and climate change which may have important implications for disentangling vaccine escape, vaccine implementation strategies, and for hospital capacity preparations in changing environments. Critically, it also highlights the need to meet the policy thresholds for PM_2.5_ to directly improve human health.

## Materials and Methods

### Data Processing

#### Pneumococcal Disease Datasets

##### Epidemiological Cases

The case counts of invasive pneumococcal disease (IPD) were provided by the National Institute for Communicable Diseases as part of national surveillance encompassing all nine provinces GERMS-SA. We included the entire dataset which comprised culture positive with viable isolates available for further analysis, culture positive with no viable isolates available, and culture negative with metadata results from PCR. Isolates were phenotypically characterised as has been previously described^7,63^. Finally, these included pneumococcal disease line list cases (n=59,017) between 2005-2023 collected from 531 hospitals across 52 districts in South Africa (Figure 1A-B). These excluded n=661 cases from the original dataset without hospital geolocation. Most cases were found in Gauteng with 24,303 cases. Northern Cape reported the least number of cases, with 1,067 (Figure S1C). Disease types included 32,639 bacteremia (55.3%), 21,167 cases of meningitis (35.9%), and 5,211 other IPD cases (8.8%), such as empyema, septic arthritis, endophthalmitis, peritonitis, or pericarditis. These were classified given specimen type whereby meningitis cases were isolated from cerebrospinal fluid specimens, bacteremia from blood specimens, and other invasive disease classes were from other typically sterile sites. Bacteremic cases do include bacteremic pneumonia but non-bacteremic pneumonia is not included. The median age was 32 with 13% of cases reported in children <1 year old. There was missing age data for 4.3% of cases and serotype information was missing for 37.8% of cases (Figure 1C). We included data from 2005-2023 in concordance with the environmental datasets for the initial model testing. As the genomic dataset ran until 2020, the year marking the beginning of the COVID-19 pandemic (with a substantial decrease in IPD counts), we restricted the models to the period 2005–2019 (n=52,437) so as to overlap with epidemiological cases and exclude this case reduction. In the overall environmental epidemiology models we included all data and for the models stratifications by outcome we included all reported data for each outcome. The outcomes we assessed included different age groups, disease types, and serotypes.

##### Genomic Sequences

The genomes included in this study were sequenced as part of the Global Pneumococcal Sequencing project, GPS^64^ which is a global genomic survey of *S. pneumoniae*. Isolates were collected by the National Institute for Communicable Disease in South Africa from 2005-2020 (n=4,912) from 48 of the 52 districts. We only include genomes through the year 2019 (n=4,350). Isolates were randomly sampled using a previously described protocol whereby approximately 300 invasive-disease isolates from each year (2005–2014) were selected with a specific target age breakdown (50% from <3 year olds, 25% from 3–5 year olds, 25% from >5 year olds). In the second phase of sequencing, 100 disease isolates from 2005 to 2010 from children <5 years old were additionally, randomly sequenced. The sampling for 2015-2020 was similar however the sampling strategy was not adjusted for the reduction in IPD cases following PCV (thus included more >5 year-olds)^6,7,65^. Most sequences are from the districts with the highest population size (Figure S18). Assembly quality control parameters included a minimum average sequencing depth of 20× and an assembly length of 1.9–2.3 Mb. Sequences with more than 220 heterozygous SNP sites were excluded. The genomes were assembled using Spades assembly software^66^ and the gene annotations were made using Prokka^67^ PopPunk^68^was used to assign each genome to a Global Pneumococcal Sequence Cluster (GPSC). We included each GPSC with an N>90 and included GPSC8 due to its public health relevance^7^. We included the proportion as the number of each GPSC over the total number of sequences per week per province (Figure S19).

##### Sociodemographic Data

The sociodemographic data was extracted from multiple sources. The population size per year per district (from 2005-2023), were sourced from the 2023 Statistics South Africa projections^69,70^From the same report we included per province ART coverage, HIV prevalence, life expectancy, growth rate, % children (<15) and the % elderly over time (60+). We also included the aging index and dependency ratio. These are defined by Statistics South Africa as the number over age 65 per 100 youths and the number <15 years old combined with the number 60+ over the number of working age population (15-64) respectively.^70^ The pneumococcal conjugate vaccine has a 2+1 schedule in South Africa and was first introduced with PCV7 in 2009 and PCV13 in 2011 (Table 1). PCV coverage for 1, 2, and 3 doses were included from the WHO Immunization Data report.^71^ Three doses is defined as the percentage of surviving infants who receive the 3rd dose of pneumococcal conjugate vaccines. We included the official coverage estimates as well as the WHO/UNICEF Estimates of National Coverage (WUENIC)^72^ (Figure S1D). The official estimates initially are based on surveys and other administrative messages, which, due to biases and inaccuracies, are adjusted by the national authorities to provide estimates of the most likely coverage.

#### Environmental Datasets

South Africa comprises temperate and arid climates according to Köppen classification^73^. Gauteng and Western Cape provinces (where the majority of our case data is from) are sub-tropical highland and Mediterranean climates respectively.

##### Meteorological Data

The environmental datasets spanned 2005-2023 inclusive. We included temperature, precipitation, relative, and absolute humidity metrics from the global reanalysis dataset ERA5-Land^74^ aggregated for weekly time points at the district level. This is gridded data with 0.1° degree resolution (accounting for the mean latitude of South Africa at -29° so each grid square is approximately 108km^2^). This resulted in a mean of 218 grid cells per district ranging from 15 in Johannesburg, Gauteng to 1180 in Namakwa, Northern Cape. Daily temperature metrics were retrieved and aggregated to weekly such that we include the weekly mean of the daily mean, maximum, or minimum temperature. Precipitation was initially retrieved as hourly average meters (m) per second precipitation loss rate (prlr). We converted it to daily millimeters (mm) and then tested inclusion of the weekly total precipitation and the mean daily precipitation.

We calculated the Standardised Precipitation and Evapotranspiration Index (SPEI) at 3-month and 6-month time periods using the above mentioned precipitation and temperature data from ERA5-Land. The packages used for this calculation were SPEI^75^, ClimProjDiags^76^, CSindicator^77^, and CSTools^78^.

Absolute humidity (absh) is defined as the mass of water vapour in a volume of air and the units are g/m^3^. Relative humidity (hurs) is the water vapour present in the air relative to the saturated air and defined as the partial pressure of water vapour as a percentage of the saturation pressure (%)(Figure 1D).

##### Air Pollution Data

The air pollution datasets spanned 2005-2023 inclusive. To determine which estimates of air pollution to include we compared air pollution estimates obtained from Copernicus Atmosphere Monitoring Service (CAMS) global reanalysis dataset, and NASA’s MERRA-2^79,80^ We performed an assessment of the correlation between the gridded air quality reanalysis models against observational data made available by the South African Air Quality Information Services (SAAQIS)^81^. PM_2.5_ and PM_10_ are not directly included in MERRA-2, so they were computed using methods detailed in the MERRA-2 documentation^82^(Figure 1D)(Table 2).

We included the MERRA-2 product from NASA for the entire time period and also included the bias corrected MERRA-2 product from 2020-2023^83^ The CAMS global datasets are at approximately an 80km^2^ resolution while the MERRA-2 datasets are at a 50km^2^ spatial resolution. We also included a gapless 1km gridded machine learning model from Wei *et al.,* 2023^84^

We extracted observational data from air quality monitoring stations provided by SAAQIS in South Africa. We calculated a mean absolute error, root mean square error, and Spearman’s correlation between the reanalysis products and the mean across observations in the district of Gert Sibande, Mpumalanga, Gauteng province, and KwaZulu-Natal province (Figure S23A). These were the locations with the most consistent observations across the time period available in SAAQIS. We performed a formal comparison of the datasets against observational data and found that CAMS was the best performing reanalysis model (Figure S23B). CAMS performed best despite the poor performance in the Gert Sibande region where none of the products performed well, either due to the observations being flawed, or a bias across all reanalysis products impacting this region. We included the air quality reanalysis data from CAMS in our study (Table 3).

The R package startR was used to retrieve and reformat all multidimensional environmental datasets^85^.

### Epidemiological Model Specification

#### Bayesian hierarchical mixed models using integrated nested Laplace approximation

In brief, here we summarise our modeling strategy for identifying meteorological and air pollution factors that drive patterns of pneumococcal disease; with a deeper analysis of the effects of PM_2.5_ on different outcomes (e.g. serotype, age group, disease type).

To assess the risk from environmental factors on IPD we adopted the following modeling strategy:

1. We first developed a sociodemographic base model which accounted for population density and vaccination period as well as residual seasonal, spatial, and interannual effects.
2. We tested different metrics for vaccination and antiretroviral therapy to assess their impact on disease patterns. We chose a sociodemographic base model given a balance between model fit and tractable complexity ultimately including confounding effects of vaccination as a categorical variable (across 3 vaccination periods), and population density as a continuous variable, alongside the residual random effects.
3. We then developed a meteorological model that utilized delayed lag non-linear models (DLNMs) to assess the risk from weekly, district level, meteorological factors (m_s,t_) across lag periods. These included absolute humidity, relative humidity, mean, minimum, and maximum temperature, precipitation, wind speed, and the Standard Precipitation Evapotranspiration Index (SPEI; a drought indicator). We evaluated model fit and effect size across 8-weeks of lag time from exposure.
4. To elucidate the risk from air pollution we developed an air pollution model that included a DLNM crossbasis for PM_2.5_, PM_10_, and SO_2_ including the same sociodemographic base model and meteorological confounding factors. We tested different formulations for inclusion of the meteorological confounding including linear and non-linear covariates of humidity and temperature with different lag-times. We also evaluated the inclusion of a temperature crossbasis alongside the PM_2.5_ crossbasis, and temperature as a modifying interaction with the PM_2.5_ crossbasis. We found the WAIC to be similar across the temperature covariate model, the temperature crossbasis model, and the interaction model. The PM_2.5_ effect sizes were also unchanged across these model types. The final air pollution model included a non-linear lag 3 humidity covariate and linear lag 0 covariate for temperature.
5. Using our final air pollution model, we then focused on PM_2.5_ to determine how the risk from PM_2.5_ varies given different outcomes (e.g. serotype, age group, disease type).
6. Finally we incorporated a modification by different underlying genomic lineages with PM_2.5_ in the same model so as to elucidate how the underlying microbial population modifies these PM_2.5_ effects. The PM_2.5_ air pollution models all accounted for the potentially confounding effects of temperature, humidity, population density, and vaccination.

#### Statistical Analysis

All statistical analysis and visualizations were conducted using R version 4.4.2^86^ with key packages including R-INLA^87^, dlnm^88^, dplyr^89^, sf^90^, and vegan^91^.

##### Sociodemographic Base Model Development

We began by building a model to account for residual spatiotemporal effects and important sociodemographic covariates such as population density, pneumococcal conjugate vaccine (PCV), or antiretroviral therapy (treatment for HIV; ART). We modeled weekly or monthly pneumococcal disease counts in South Africa between 2005-2023 or for 2005-2019 (excluding the post-COVID-19 period), by specifying a Bayesian hierarchical mixed model using pneumococcal disease case counts at the district (n=52) or province (n=9) level as the response variable. To account for overdispersion, we assumed that case counts, y_s,t_ where *s* represents a spatial unit and *t* represents time step (1 to 987 weeks, or 228 months) following a negative binomial distribution such that:

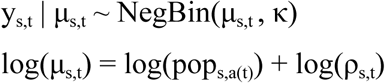

where μ_s,t_is the expected number of cases and κ is the overdispersion parameter. The distribution mean, μ_s,t_, is defined as the product of the population, *pop_s,a(t),_* (per 100,000 people) in a given spatial unit (district or province), *s*, and year, *a(t)*, with the disease incidence rate ρ_s,t_ estimated via a combination of covariates and random effects.

For all sociodemographic model formulations we evaluated different spatial (district or province) and temporal (weekly or monthly) resolutions ultimately only including the weekly district level models in our main analysis (Table S2).

###### Spatiotemporal random effect assessment

We evaluated model performance defining the response variable both at district and province level for weekly and monthly case counts and across various random and fixed effect combinations comparing them to the intercept only model (Table S2). These models included an intercept α as well as temporal random effects accounting for unknown and unmeasured monthly variation (calendar months January (1) to December (12)), δ_m(t)_, and interannual effect γ_a(t)_at the national level. We tested both the monthly variation, δ_m(pr(s),t),_ and interannual effect, δ_m(pr(s),t),_ replicated by province *pr(s)*, (n=9 provinces). We also included spatial effects with a Besag-York-Mollié (BYM-2)^92^ with both structured (*u_s_*) and unstructured effects (*v_s_*) at the district and province level model. In the province level model *s* = *pr*(*s*). We additionally evaluated including the spatial random effects with a yearly replication to adjust for fine-scale spatiotemporal confounding (Figure S13).

###### Impact of pneumococcal vaccine and antiretroviral therapy on interannual variation

We assessed different formulations of pneumococcal vaccination, antiretroviral therapy, and population density. We tested inclusion a single categorical covariate with three-phases to account for decreasing case counts after the implementation of the PCV (*PCV_periods_*): pre-PCV7 (years before 2009), PCV7 (2009-2010 inclusive), and PCV13 (2011 onwards). We alternatively assessed models including only two periods for PCV (*PCV_2009Vaccines_*): pre-PCV (years before 2009; as above), and then a single post-PCV period (2010 onwards). We also evaluated models with national level PCV coverage (%) per year (*PCV_coverage,a(t)_*), as well as a model accounting for two time periods for COVID-19 (*covid*): pre-covid (2005-2019) and post-covid (2020 onwards). To assess the impact of the uptake of ART across South Africa we included a variable for ART coverage. These included a national level ART coverage (%) (*ART_coverage,a(t)_*) and province level ART coverage (%) (*ART_coverage,pr(s),a(t)_*).We tested inclusion of the intervention variables (*PCV*, *ART*, *covid*) independently and together where *f* indicates a non-linear function. We also included district level human population density (*pd*) as a continuous variable due to its particular relevance as a confounding factor with air pollution.

In summary the main models tested included: i) PCV as a categorical variable with 2 categories i) PCV_2009Vaccines_ (M10), ii) PCV_periods_ (M11), iii) PCV_coverage,a(t)_ (M13); iv) ART_coverage,a(t)_ (M14); v) ART_coverage,pr(s),a(t)_ (M15); and vi) ART_coverage,pr(s),a(t)_ and PCV_periods_ (M16) (Table 4). M12 is the same as M11 with additional inclusion of population density. These were all run with and without a replication of the interannual effect across provinces (Table S4). We also ran different models for different types of IPD to assess the impact of these perturbations across different groups. These included children ≤5 years old, adults between 15 and 64 years old, adults 65 and above, IPD from only NVTs and from only VTs. We also ran these models including disease from 2005-2019 (excluding the post-COVID-19 period), and from 2005-2023 (Table S4).

We found an increasing goodness-of-fit with increasing complexity (Table S2). We included M12 as our sociodemographic base model due to the balance of complex random effects and goodness-of-fit. This model was formulated such that (Equation 1):

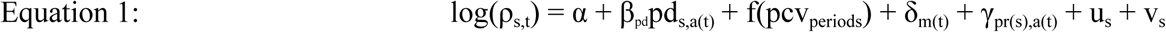

#### Distributed lag non-linear models to estimate disease risk with environmental factors

We included models with the above described sociodemographic base model (Equation 1). We evaluated the Pearson correlation between all environmental variables. Then to explore the relationship between exposure lag-times and disease, we correlated lag times from exposure to disease outcome using a Spearman correlation (Figure S3). We evaluated IPD risk from meteorological factors and then air pollutants, while controlling for confounding meteorological factors. We tested all models including both the province of Gauteng alone and excluding Gauteng province; due to it being the most populous, highest incidence of IPD, and highest maximum concentrations of PM_2.5_, PM_10_, and SO_2_ (Figure S8)(Figure S9)(Figure S10)(Figure S11). While we evaluated many models across different spatial and temporal scales, our final meteorological and air pollution models presented in the main manuscript were at the weekly district level from 2005-2019 (Table S5). We estimated the exposure-lag-response association using the crosspred() of the dlnm package for all DLNMs to generate estimates and Gaussian-approximate credible intervals derived from the INLA posterior confidence intervals. Although crosspred() typically reports confidence intervals, in this case the uncertainty reflects the Bayesian posterior distribution because the supplied coefficients and covariance matrix were obtained from INLA.

##### Meteorological models

We developed a meteorological model to assess the effect of meteorological factors on pneumococcal disease dynamics. We added DLNMs for all meteorological factors to the sociodemographic base model (Equation 2). These included temperature mean (tas), minimum (tasmin), maximum (tasmax), absolute (absh) and relative humidity (hurs), precipitation mean (prlrmean) and sum (prlrsum), Standardized Precipitation Evapotranspiration Index (SPEI; drought indicator) for 3 months (SPEI3) and 6 months (SPEI6), as well as windspeed (sfcWind). To account for lag times from the exposure to disease outcome we specified distributed lag non-linear models (DLNMs) to estimate risk of pneumococcal disease incidence for different environmental factors and lag timeS10^88,93^. We included cases from 2005-2019 to match our genomic sequence models. We ran weekly models at both province (adm1) and district (adm2) level. We evaluated 6, 8, and 12 week lag times across the variables and administrative regions finding little difference across the cumulative effects for each model., We proceeded with models including lags of up to 8 weeks from environmental exposure to pneumococcal disease due to our own model comparison and previous work identifying effects up to 4 weeks after environmental exposure^18,34^. Furthermore we found the maximum correlation between the disease outcome and environmental exposure up to around a 5-week (Figure S3). Given the statistical evidence together with the estimated 30 day generation time for the pneumococcus, an 8-week lag period is expected to capture both the statistically and biologically relevant effects^6^. We generated the DLNM crossbasis with 8 weeks of lag with 2 degrees of freedom for the lag knots except for relative humidity for which we included 3 degrees of freedom to capture the complex non-linearity. For each variable we included equally distributed knots at the 33rd and 67th quantile except for precipitation for which we included 2 equally spaced knots on a log scale of the precipitation distribution. We centered and scaled all meteorological factors. We centered the effect estimation at the median of the variable distribution (Table S5).

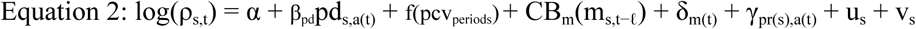

##### Air pollution models

We developed an air pollution model to assess the risk from air pollution on IPD. We included the same model as above with vaccination period and population density covariates as well as residual spatiotemporal effects and then tested different formulations of confounding meteorological effects. Meteorological factors are known confounders of air pollution and their inclusion also ensures that our model is applicable across varied climates. We developed our meteorological covariates given the known interactions between temperature and air pollution, and humidity and temperature^92^. Our final air pollution model included temperature as a linear term with no lag-time and absolute humidity as a non-linear term with a 3-week lag-time (Equation 3). We chose this formulation given their effect sizes at these lag times, and comparison against other model formulations for the different confounders (described below) (Figure S8)(Table S5).

We assessed air pollution models with the confounding effects of mean or maximum temperature alone, as well as with the additional effects of absolute or relative humidity. We formulated the humidity and temperature models including both variables at no-lag time, a 3-week lag time, or a mixture of one at 3-weeks, and the other at 0. We included the humidity variables as non-linear effects using random walk 2 models with 5 cuts and included the temperature variables as linear effects given their exposure response curves from the meteorological models (Figure 4)(Figure S5). We also assessed directly interacting a PM_2.5_ crossbasis with a mean temperature covariate, as well as including an additional mean temperature cross basis together with absolute humidity covariate, finding similar goodness-of-fit and risk estimates across all different formulations including both humidity and temperature (ΔWAIC <4)(Table 5)(Table S5). For parsimony, and given the goodness-of-fit metrics we proceeded with the covariate model including lag-week 3 absolute humidity and mean temperature covariate with no lag-time (Equation 3).

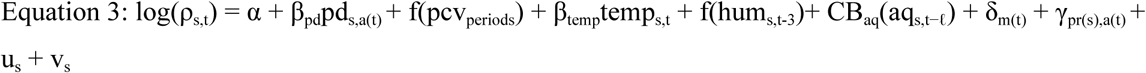

We centered the air quality metrics at the 24-hour National Ambient Air Quality Standards (NAAQS) for South Africa (PM_2.5_ = 40 μg/m^3^, PM_10_ = 75 μg/m^3^, and SO_2_ = 125 μg/m^3^). We utilized the 24 hour exceedance threshold rather than the annual as we are examining an acute IPD effect (Table 6)(Table S5).

**Table 6.**
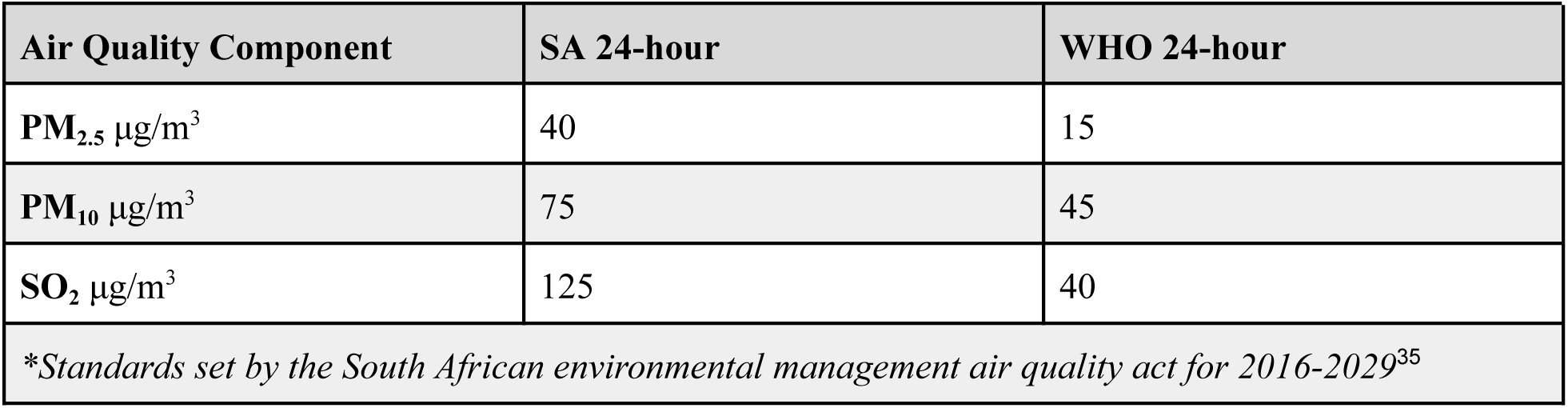
Air quality standards. For the years 2016-2029 as set by the Environmental Management Air Quality Act 39 in South Africa and the standards set by the WHO in 2021.

| Air Quality Component | SA 24-hour | WHO 24-hour |
| --- | --- | --- |
| $PM_{2.5} \mu g/m^3$ | 40 | 15 |
| $PM_{10} \mu g/m^3$ | 75 | 45 |
| $SO_2 \mu g/m^3$ | 125 | 40 |
| *\*Standards set by the South African environmental management air quality act for 2016-2029<sup>35</sup>* |  |  |

##### PM_2.5_ effect across age, disease, and vaccine status

To identify differences in risk from PM_2.5_ across different stratification groups we used our air pollution model formulation but changed the outcome from IPD overall to disease incidence per 100,000 people among each group for age group, disease outcome, vaccine status (whether cases were serotypes which were included in the PCV7 vaccine, PCV13 vaccine, or were non-vaccine type), and by specific serotype(Table 4). The specific models we tested included i) age ≤5 years old, ii) age 6-14 years old, iii) age 15-64 years old, iv) age ≥65, v) bacteremia, vi) meningitis, vii) other invasive disease types, and viii) penicillin resistant cases (measured at the meningitis threshold). We also included serotype stratifications by ix) NVT, x) PCV7, and xi) PCV13. Models xii-xxix included stratification by IPD from particular serotypes (Table S1). To estimate the relative risk ratio (RR ratio) between each subgroup (serotype, disease type, age group) compared to the overall population we crosscut the relative risk from the exposure-response curve at 50 μg/m^3^ concentration for each. We then included the relative risk for each subgroup in turn as the numerator compared to the relative risk from the overall model to estimate the increased (>1) or decreased (<1) risk of IPD, in each subgroup, from PM_2.5_ compared to the risk for pneumococcal disease overall (Figure 4A).

#### Distributed lag non-linear models interacting with lineage prevalence

To assess the impact of underlying lineage diversity on the environmental effects we utilized the same air pollution model framework as above (comprises sociodemographic base model (M12: seasonal, spatial, and province-replicated-interannual random effects, together with vaccination periods and population density, and adjustment for temperature and humidity)(Table 4). We included all GPSCs with N≥100 as well as GPSC8 due to its epidemiological importance in South Africa^6,65^. We then included an interaction between the exposure crossbasis and a modifying variable representing GPSC-specific proportions per total number sequenced each week of dominant GPSCs. We ran these models at the province level weekly and thus included a province level spatial effect for these interaction models. To ensure model interpretability, we scaled and centered this variable (Table S6).

We evaluated the exposure–response relationship at three representative values of the effect modifier, corresponding to the 0.1, 0.5, and 0.8 quantiles of the centered distribution of the modifying variable (excluding zeros). For each quantile value *z*, we derived level-specific coefficient vectors and corresponding variance–covariance matrices that reflect the combined main and interaction effects.

To account for uncertainty in the interaction effects across different levels of the modifying variable, we computed level-specific coefficient vectors and their corresponding variance-covariance matrices. For a given value z of the centered interaction variable, the coefficient vector was calculated as:

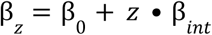

and the associated variance-covariance matrix as:

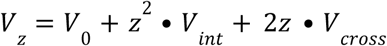

where β_0_ and *V*_0_ are the estimated coefficients and variance-covariance matrix for the main (non-interaction) effect, β_*int*_ and *V*_*int*_ are those for the interaction term, and *V*_*cross*_ represents the covariance between main and interaction coefficients. These were then passed to the crosspred() function of the DLNM package to generate estimates and Gaussian-approximate credible intervals derived from the INLA posterior at each value of *z* (Table S6).

We performed a sensitivity analysis whereby for GPSC pairs, which share the same dominant serotypes, we split the dataset into the pre- and post-vaccine period to ensure the effect was not just driven by serotype replacement in the time series but by some underlying evolutionary process (Figure S21).

To estimate goodness-of-fit for each model we calculated the Watanabe-Akaike information criterion (WAIC)^95^and the mean absolute error (MAE). We also calculated the sum of the log10 conditional predictive ordinate (CPO) for each observation to evaluate the model’s out-of-sample predictive performance. We additionally computed a pseudo-R^2^, defined as the proportionate reduction in deviance compared to an intercept-only model^96^. This measured the ability of each model to account for variation in disease incidence (0-1) whereby a model with R^2^ = 1 would indicate a model accounting for all of the variation in the data. This helped evaluate the improvement in model fit achieved by including additional covariates.

## Data Availability

All code is available in the GitHub repository (https://github.com/sophbel/env_sa_manuscript) and the associated datasets are on Zenodo.

## Authors’ contributions

SB and RL conceptualized the study. SB, CL, JK, DL, CC, SL, SM, MdP, AvG contributed to data curation and harmonization. SB, CL, JK, DL, CC, SM, MdP, GM, AvG, RL were responsible for study design. CL, JK, SM, MdP, AvG provided expert knowledge and insight from the study location. SB, GM, RL contributed to methodological development. SB did the formal analysis. SB wrote the draft manuscript.

All authors edited, reviewed, and approved the final manuscript.

## Funding Acknowledgements

This work was supported in part by a Fogarty International Center Global Infectious Disease research training grant, National Institutes of Health, to the University of Pittsburgh and National Institute for Communicable Diseases (D43TW011255). Genome sequencing was funded by the Gates Foundation (grant code OPP1189062) and Wellcome Trust (Grant reference 206194) through the Global Pneumococcal Sequencing project, and the SEQAFRICA project which was funded by the Department of Health and Social Care’s Fleming Fund using UK Aid. The views expressed in this publication are those of the authors and not necessarily those of the UK Department of Health and Social Care or its Management Agent, Mott MacDonald.

S.B. is supported by Schmidt Science Fellows, in partnership with the Rhodes Trust. C.C. and D.L. is supported by their Barcelona Supercomputing Center AI4Science Fellowship funded by the Recovery and Resilience Mechanism-Next Generation as part of the Spanish Ministry’s Recovery, Transformation and Resilience Plan (C005/24-EDCV1). R.L. acknowledges support from the Wellcome Trust (IDExtremes 226069/Z/22/Z), the European Union’s Horizon Europe research and innovation program (IDAlert, grant agreement 101057554) and a Royal Society Dorothy Hodgkin Fellowship.

In addition, we would like to thank the CRDM surveillance team at the NICD for assisting with retrieval of isolates, phenotypic characterization, managing and cleaning of the GERMS database and the Sequencing Core Facility at the NICD for the generation of additional genomic data. We would also like to thank the Atmospheric Composition group at the Barcelona Supercomputing Center for assistance with atmospheric dataset acquisition and processing. We would also like to thank Professor Rebecca Garland at the University of Pretoria, South Africa, for useful discussions regarding the performance of global reanalysis models in South Africa. We would also like to thank Dr. Carles Milà and Chloe Fletcher for helpful discussions regarding meteorological confounders in air pollution models and random effect structure.

For the purpose of Open Access, the author has applied a CC-BY public copyright licence to any Author Accepted Manuscript version arising from this submission.

