## Supplementary Figures for "Microbial diversity modifies the impact of air pollution on pneumococcal disease risk"

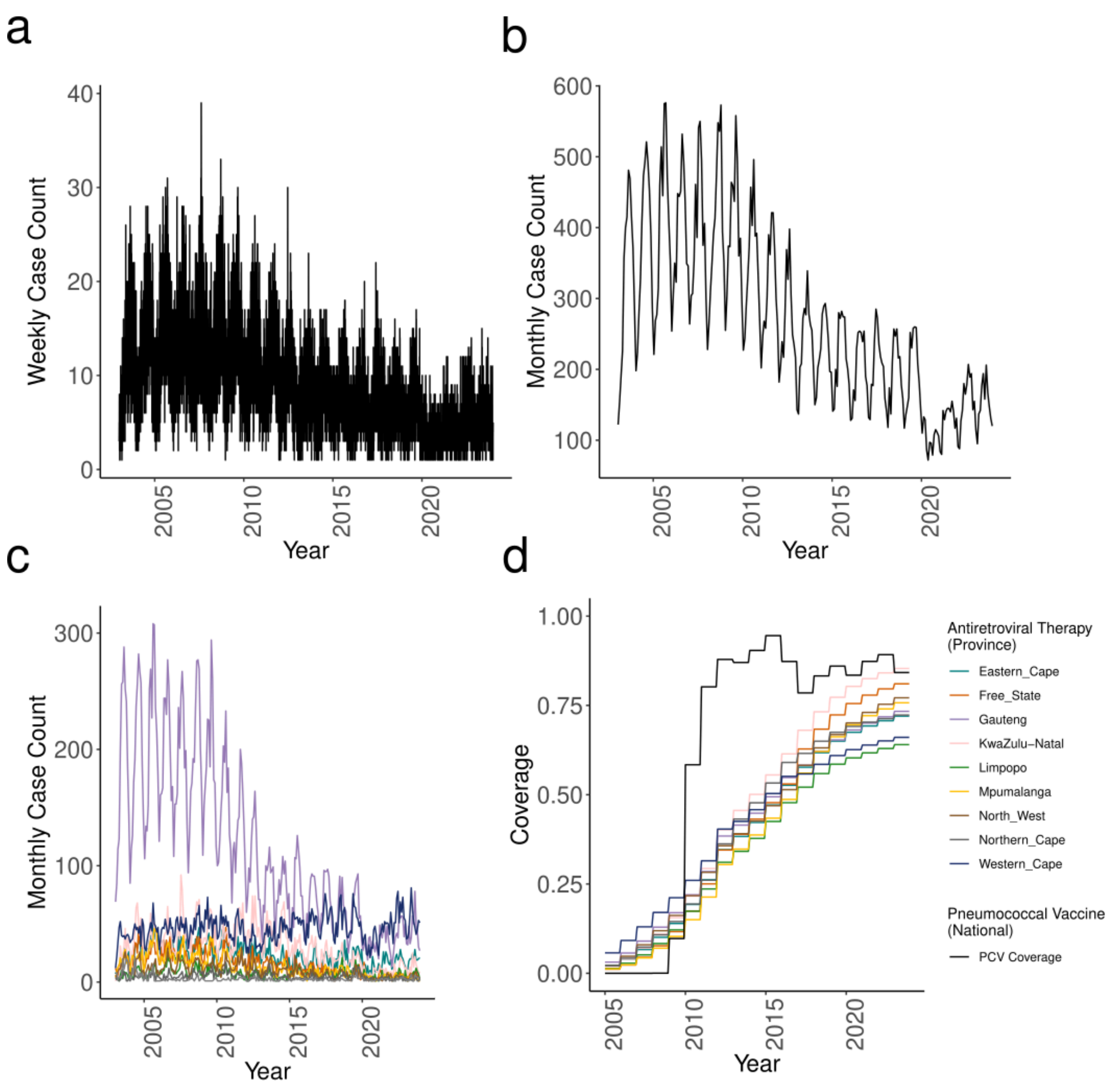


**Figure S1. Pneumococcal case counts** (a-c) and intervention coverage (d) (a) Weekly (a) and Monthly (b) pneumococcal disease case counts. (b) Disease counts monthly colored by province. (c) pneumococcal conjugate vaccine (PCV) coverage nationally and antiretroviral therapy coverage by province over time.

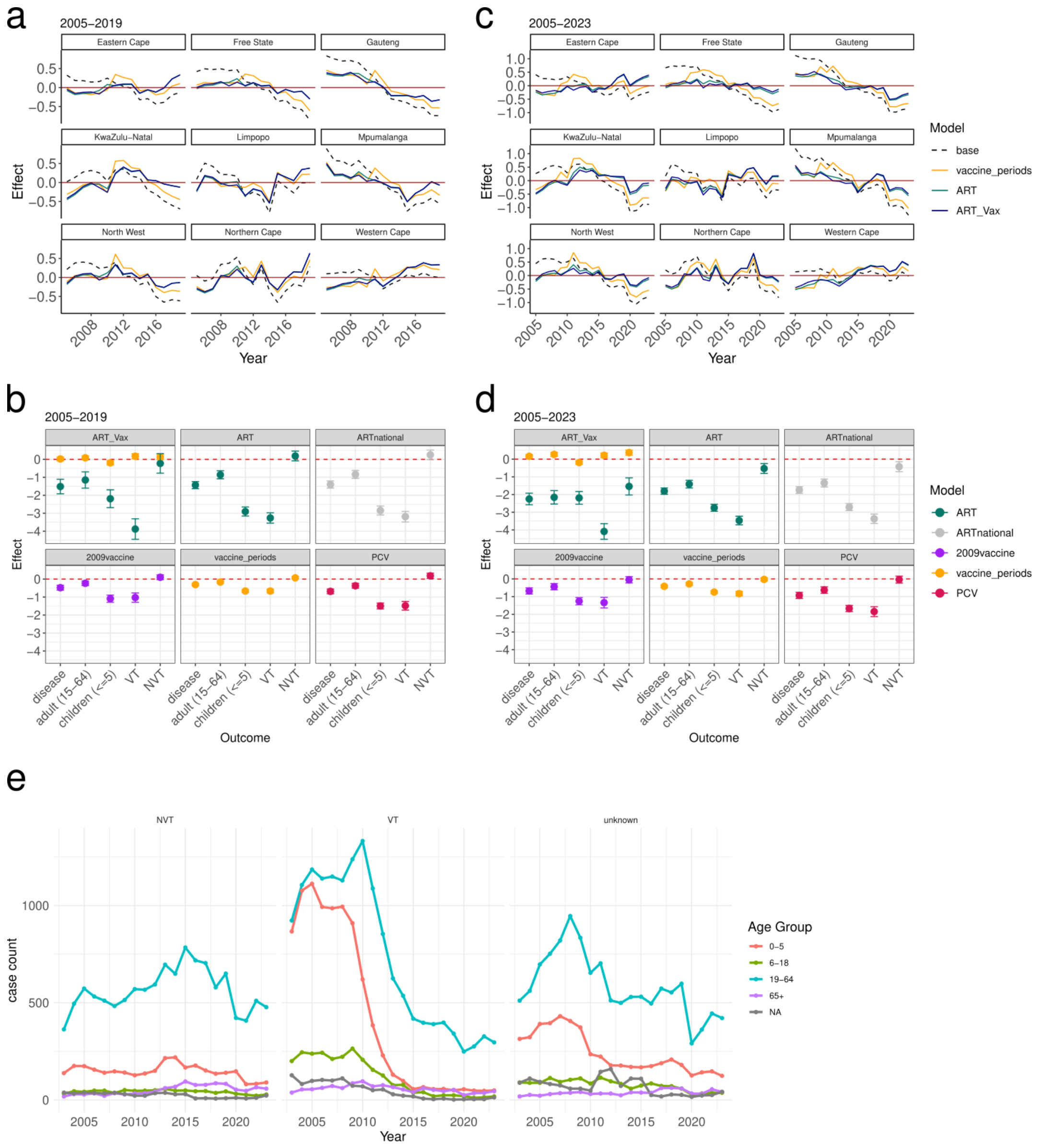


**Figure S2 (Extended Data Figure 1). Effects from implementation of ART and vaccine included in the models** with years (a-b) 2005-2019 and (c-d) 2005-2023. (a) and (c) show the interannual random effects iwith no perturbations (black, dashed), for a model with vaccination periods 2009 and 2011 (yellow), for ART coverage per province (green), and for a model including both vaccination periods and ART coverage per province (blue). (b) and (d) include a model with ART_Vax: both ART coverage per province (green) and vaccination periods (yellow), ART: a model with ART coverage per province (green), ARTnational: ART coverage nationally (grey), 2009vaccine: a model with a categorical variable indicating pre-PCV7 (2005-2008) and post-PCV (2009-2023) (purple), vaccine_periods: a categorical covariate indicating the pre-PCV, PCV7 (2009-2010), and PCV13 (2011-2023) periods (yellow), and a model with PCV coverage nationally (pink). These each were run for outcomes of all IPD, IPD in adults 15-64, children ≤5, VT serotypes, and NVT serotypes independently. (e) shows the case count across years and by age group including 0-5 (pink), 6-18 (green), 18-64 (blue), 65+ (purple), and absent age data (NA) (grey) for disease from NVTs (left), VTs (middle), and unknown serotype (right).

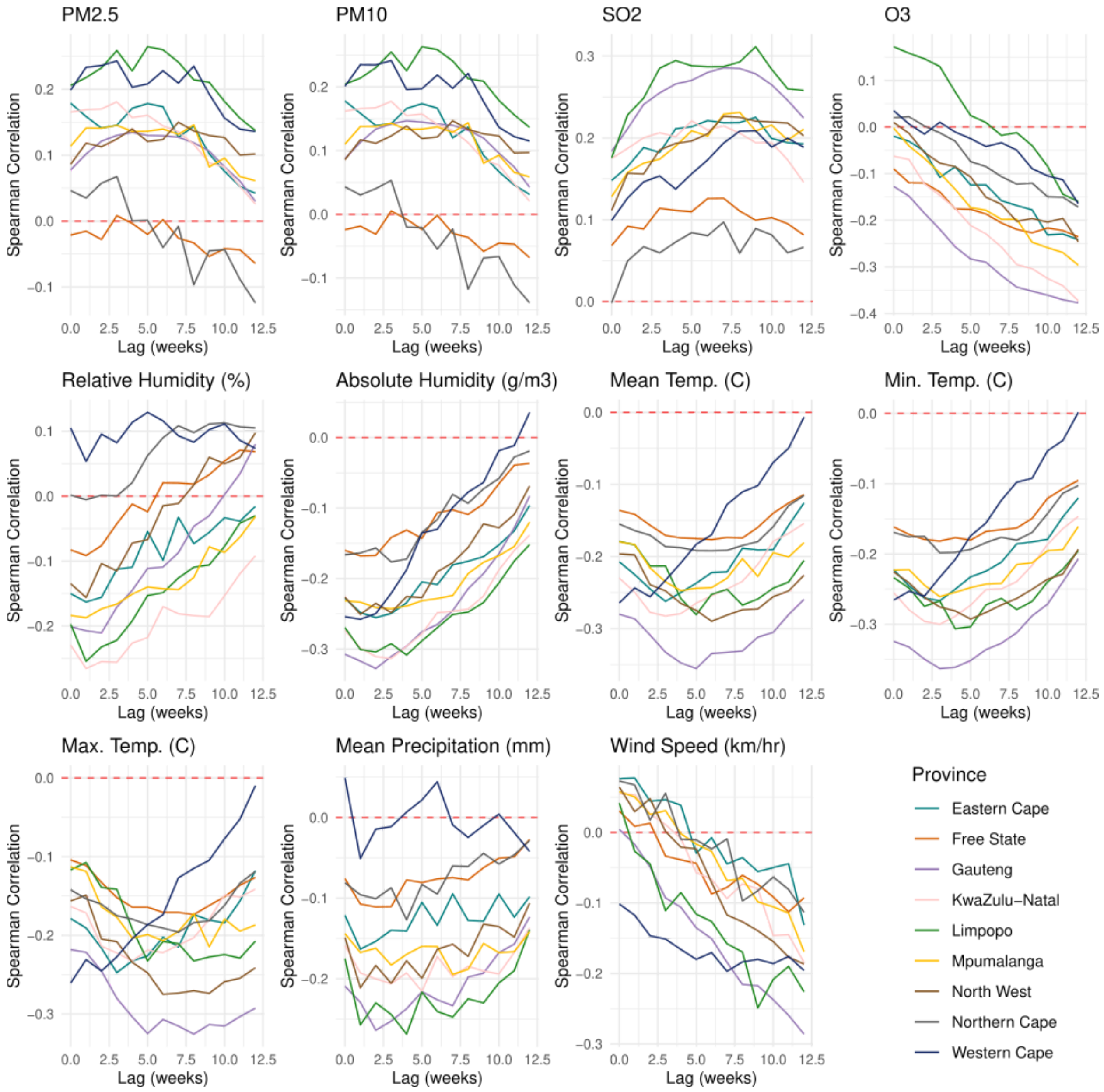


**Figure S3 (Extended Data Figure 2). Spearman correlation between each lagged variable and invasive pneumococcal disease case counts** across lag weeks with disease case counts for all environmental variables colored by province across 12 weeks of lag.

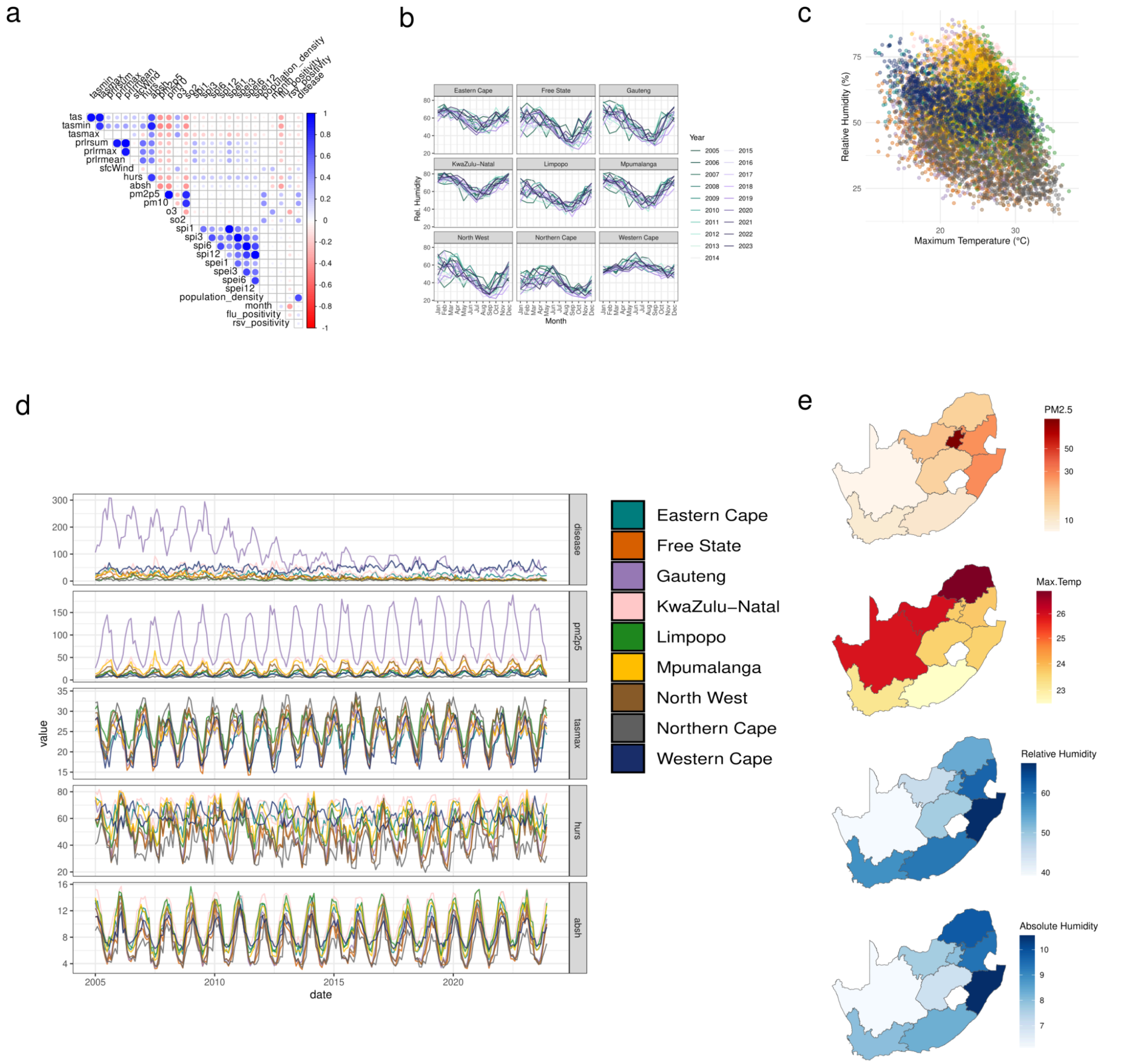

**Figure S4. Summaries of included environmental variables.** (a) correlation plot with spearman correlation between each of the external variables included. (b) relative humidity across months colored by the years of the study faceted by province. (c) Correlation between maximum temperature and relative humidity colored by province (same legend as d). (d) time series of pneumococcal disease case counts and (e) mean of each environmental variable per the 9 provinces of South Africa (d-e) $PM_{2.5}$ concentration ($\mu g/m^3$), maximum temperature (celsius), relative humidity (%), and absolute humidity ($g/m^3$ water) across 19 years colored by province.

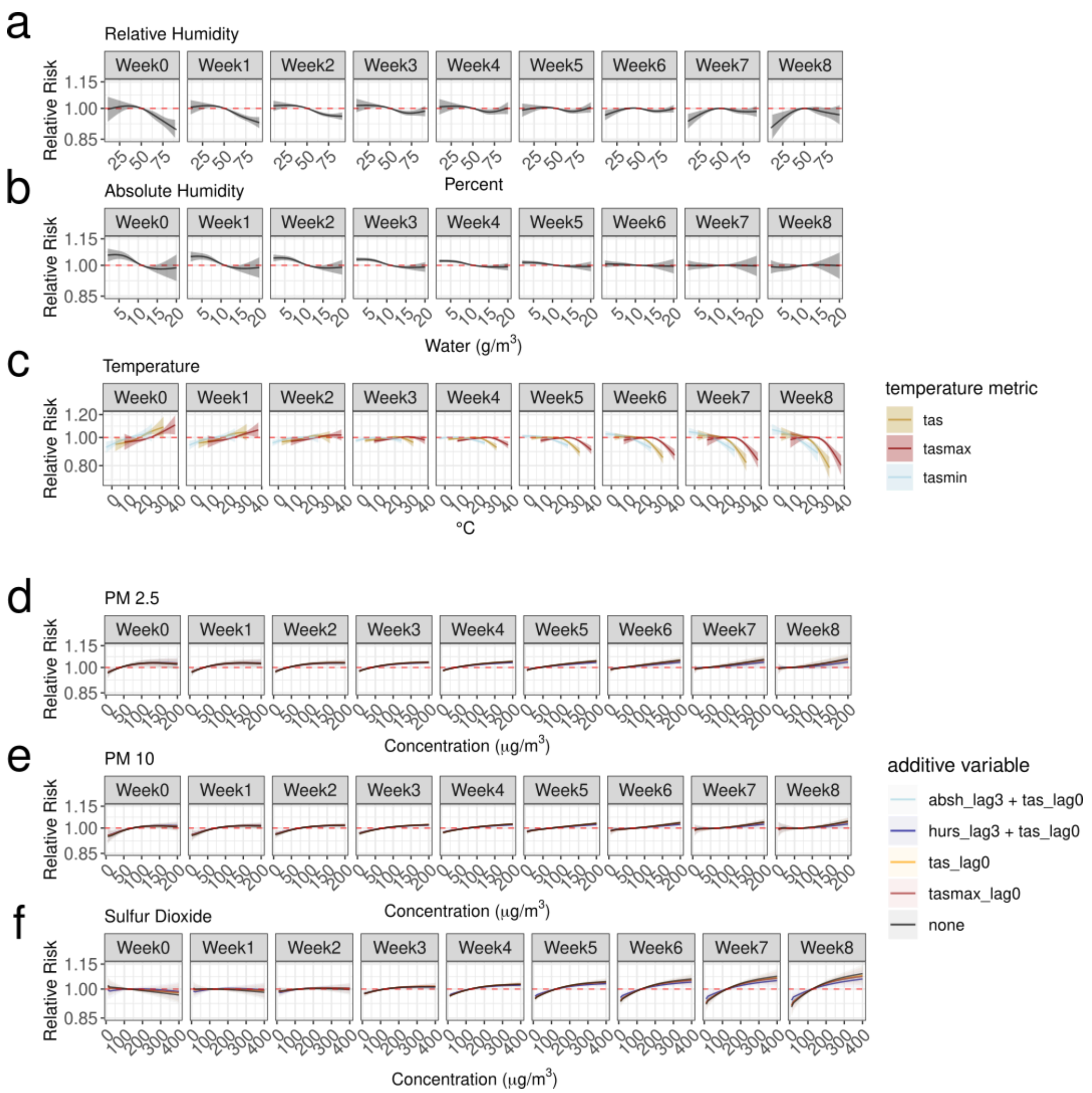


**Figure S5 (Extended Data Figure 3). Relative risk across 8-weeks of lags for relative, and absolute humidity, temperature, and air pollutants $PM_{2.5}$, $PM_{10}$ $SO_2$, relative, and absolute humidity with additional multiplicative variables.in models which account for vaccination period and population density, as well as residual spatial, seasonal, and interannual random effects.** a) Exposure response curves for (a) absolute humidity ($g/m^3$) and (b) % relative humidity. (c) Exposure response curves for maximum (pink), mean (yellow), and minimum (blue) temperature in celsius (℃). Exposure response curves for (d) $PM_{2.5}$ , (e) $PM_{10}$, or (f) sulfur dioxide including models with combinations of meteorological covariates: maximum temperature (red), mean temperature (yellow), absolute humidity and mean temperature (light blue), relative humidity and mean temperature (dark blue), and none (black). These models are fit to weekly district level data from 2005-2019 across South Africa. In the DLNM the humidity variable cross basis has 3-degrees of freedom (df), while the pollution and temperature variables have 2. The additional multiplicative effects are included as fixed effects except the humidity variables which are non-linear with 5 cuts.

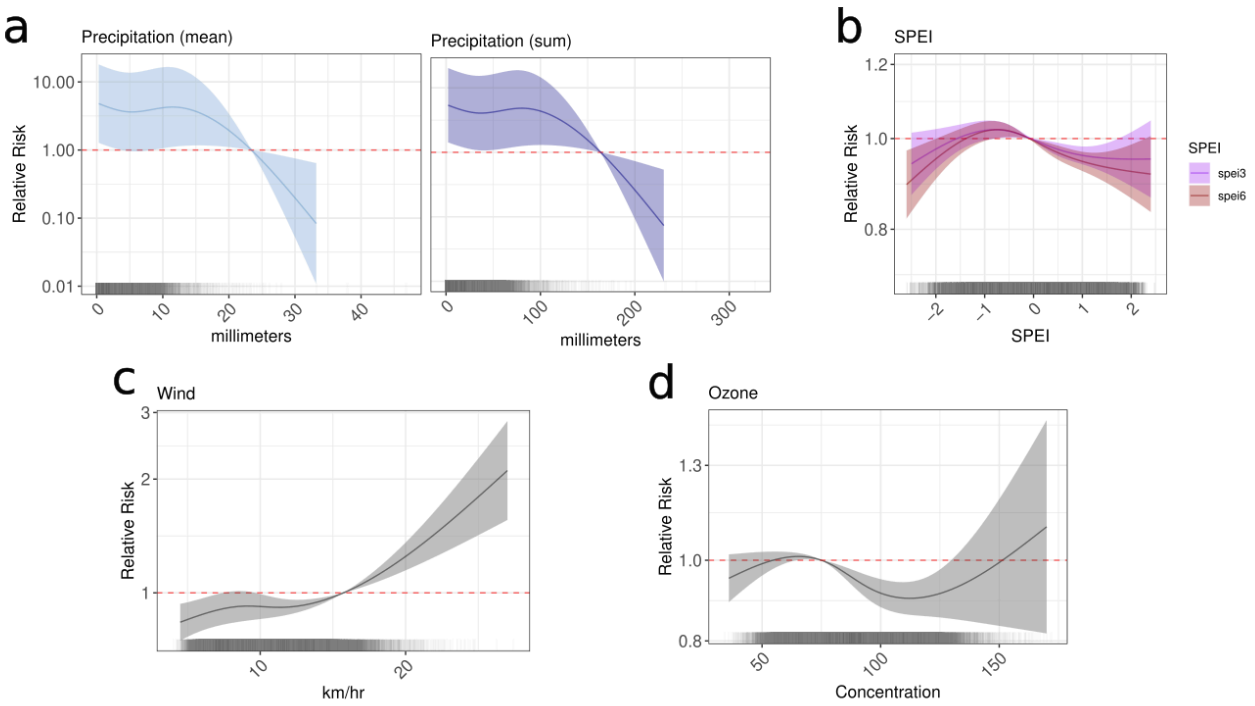


**Figure S6 (Extended Data Figure 4). Cumulative 8-week relative risk with models fit to weekly district level IPD case data** with the sociodemographic base model and these meteorological DLNMs for (a) precipitation mean (left; light blue) precipitation sum (right; dark blue), and (b) standard precipitation evapotranspiration index (SPEI) 3 months (purple) and 6 months (brown) (c) wind kilometers per hour (km/hr) (d) ozone (μg/m$^3$).

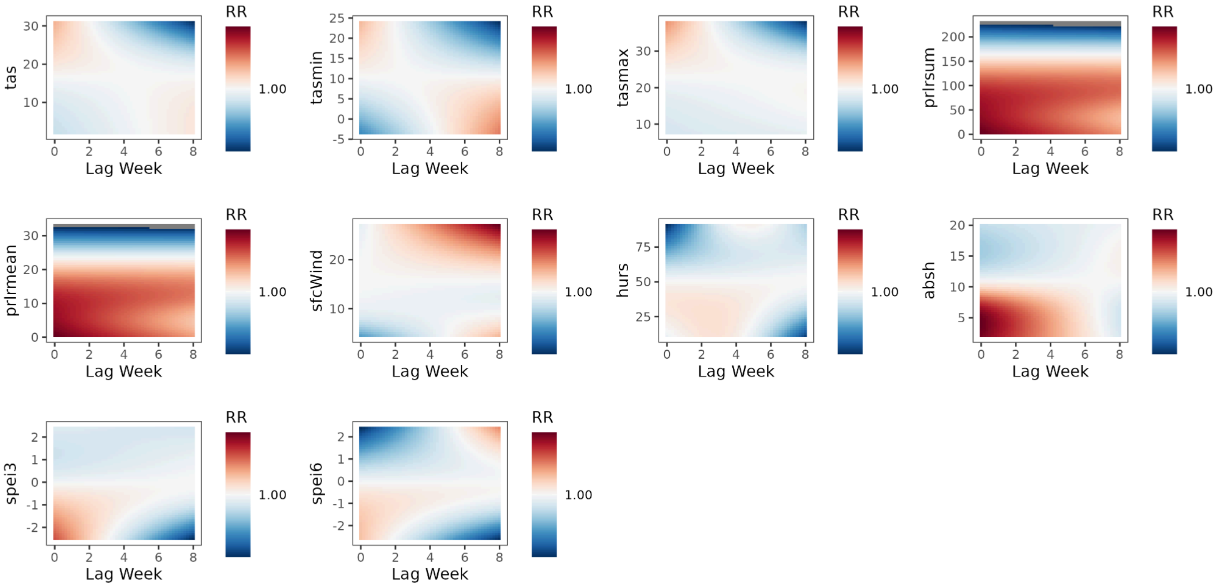


**Figure S7. Mean relative risk (RR) surfaces from meteorological models** where mean risk >1 is red and <1 is blue for temperature (celsius; mean = tas, maximum = tasmax, minimum = tasmin), precipitation (millimeters; sum = prlrsum, mean = prlrmean), wind speed (km/hour; sfcWind), relative humidity (% hurs), absolute humidity (g/m$^3$; absh), standard precipitation evapotranspiration index (3 months = spei3, 6 months = spei6).

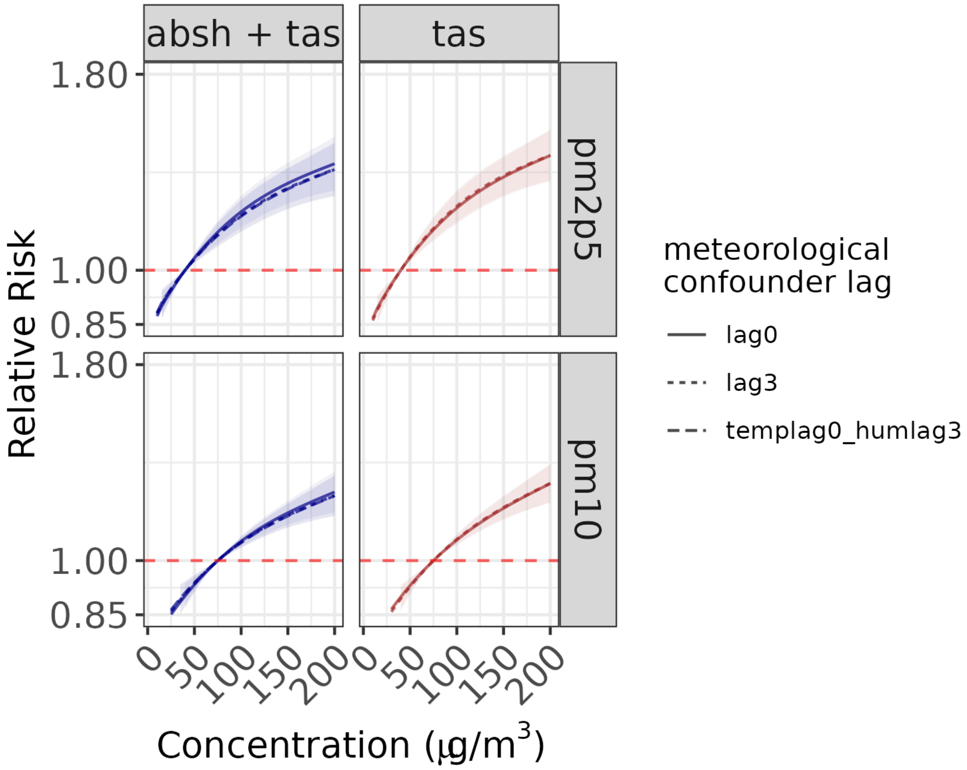


**Figure S8. Exposure-response curves across 8 weeks of lags for the relative risk** (y-axis) from each $PM_{2.5}$ (top) and $PM_{10}$ (bottom) (x-axis) in DLNM models. These models are fit to weekly district level case data with the sociodemographic base model and $PM_{2.5}$ or $PM_{10}$ DLNMs with additional absolute humidity (absh) and mean temperature (tas) incorporating meteorological confounding covariates at different lag times. Lag times incorporated here include mean temperature with no lag time (solid) and a 3-week lag (dotted) time and absolute humidity with temperature both with no lag time, both at a 3-week lag time, and with humidity at a 3-week lag time and mean temperature with no lag time (dashed). The best model formulation is the one with the mixture of humidity at 3 weeks and temperature with no lag.

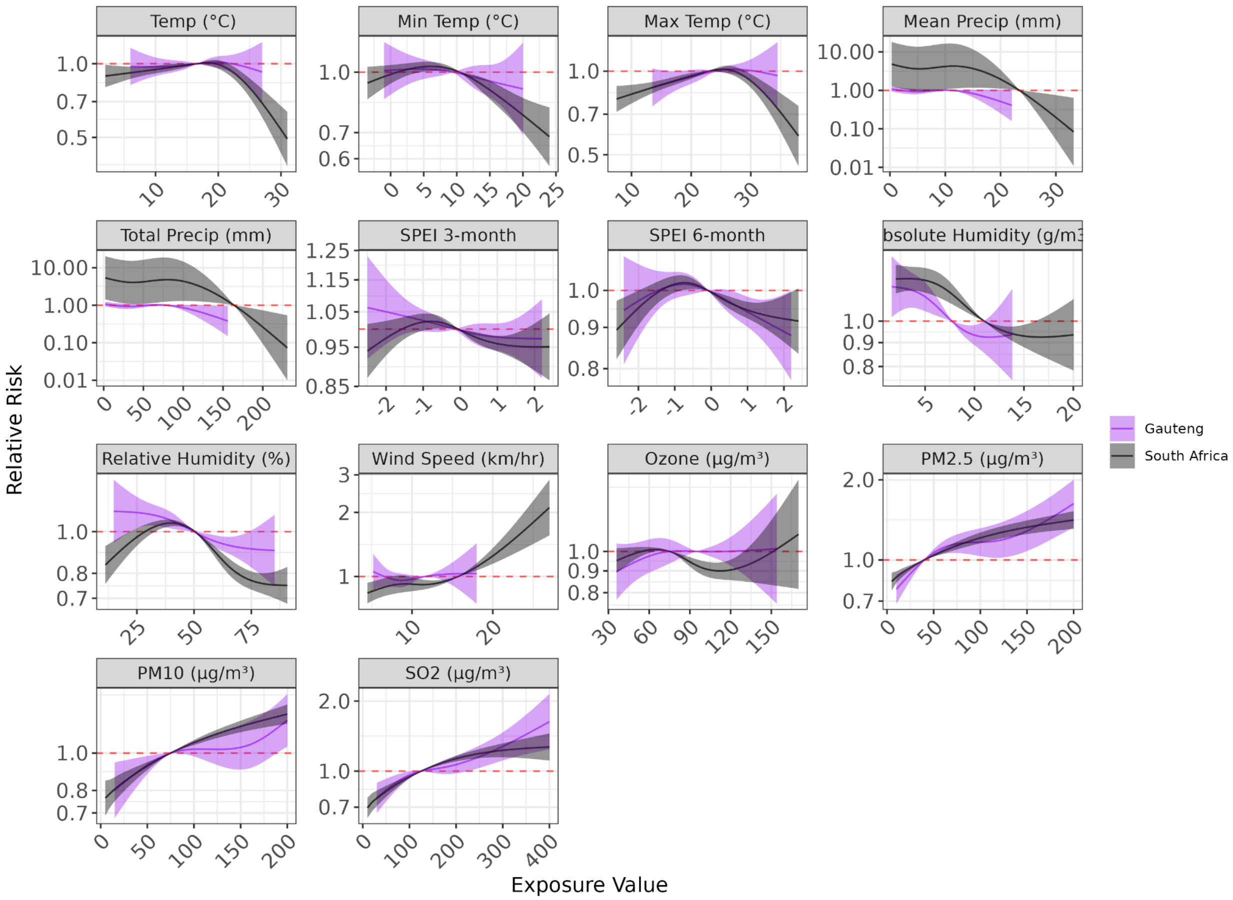


**Figure S9. Cumulative exposure-response curves across 8 weeks of lags** for the relative risk (y-axis) from each environmental variable (x-axis) in DLNM models for Gauteng alone (purple) and South Africa overall (black). These models are fit to weekly district level case data including the sociodemographic base model and distributed lag non-linear models (DLNMs) for each variable. Included variables in order are temperature (mean, minimum, and maximum celsius), precipitation (weekly mean and sum total millimeters (mm)), SPEI 3-month and 6-month, Humidity (absolute and relative), wind speed (km/hr), and ozone. For $PM_{2.5,}$ $PM_{10}$, and $SO_2$ confounding covariates mean temperature and absolute humidity are included. (a) absolute humidity (absh) in g/m$^3$ water (b) percent relative humidity (c) ozone (o3) (d) particulate matter <10μm (pm10) (e) particulate matter <2.5μm (pm2p5) (f) mean precipitation millimeters (g) cumulative precipitation in millimeters (h) mean wind speed in km/hour (i) sulfur dioxide (so2) (j) drought index including evapotranspiration across previous 3 months (k) drought index including evapotranspiration across previous 6 months (l) maximum temperature (celsius) (m) minimum temperature (celsius).

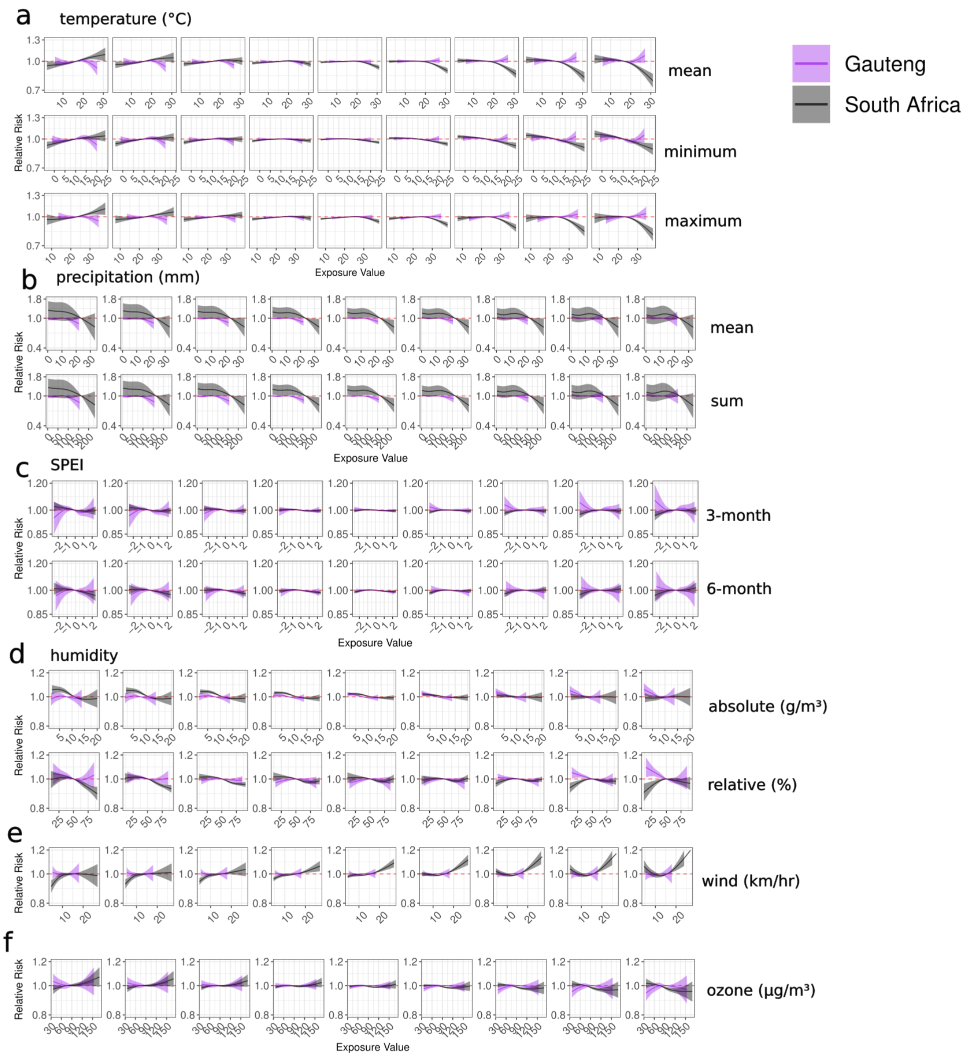


**Figure S10. Exposure-response curves across 8 weeks of lags for the relative risk** (y-axis) from each environmental variable (x-axis) in DLNM models for Gauteng alone (purple) and South Africa overall (black). These models are fit to weekly district level case data with the sociodemographic base model and additional environmental variable DLNMs. The metric for each exposure value is on the x-axis and the relative risk is on the y-axis. (a) mean (top), maximum (middle), and minimum (bottom) temperature in celsius. (b) millimeters precipitation mean (top) and sum (bottom) (millimeters; mm). (c) Standard precipitation evapotranspiration index (SPEI) for 3-months (top) and wind (kilometers per hour; km/hr), and (f) concentration of ozone (µg/$m^3$).

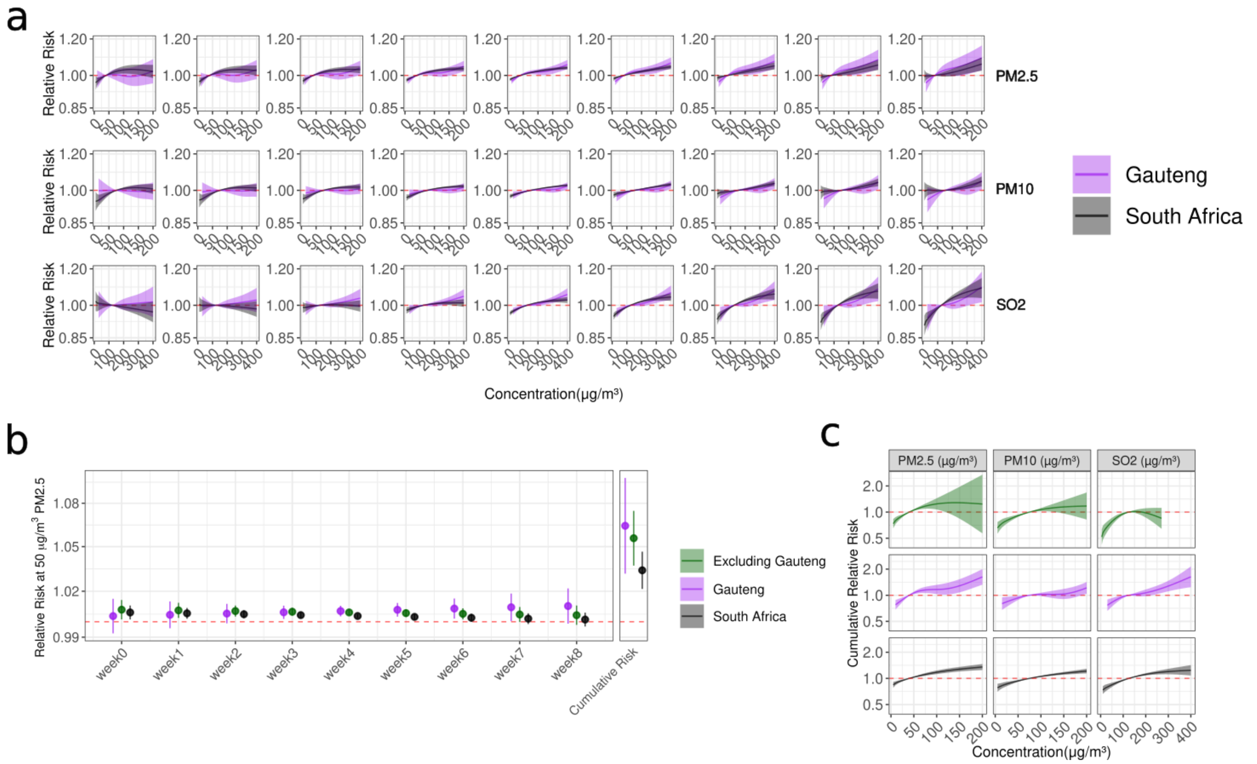


**Figure S11 (Extended Data Figure 5). Comparison of models run on all of South Africa (black), Gauteng alone (purple) and for b-c excluding Gauteng (green).** These models are fit to weekly district level case data with air pollution model (including the sociodemographic base model, temperature and humidity confounders and the DLNM crossbasis for each pollutant)**.** (a) exposure-response curves across 8 weeks of lags for relative risk from air pollutant exposure. for Gauteng alone (purple) and South Africa overall (black) for $PM_{2.5}$ (top), $PM_{10}$ (middle), and $SO_2$ (bottom). (b) Relative risk at 50µg/m$^3$ of $PM_{2.5}$ for each of the lag weeks from 0-8 and the cumulative risk. (c) The cumulative relative risk for each of $PM_{2.5}$ (left), $PM_{10}$ (middle), and $SO_2$ (right) excluding Gauteng (green), Gauteng alone (purple), and for South Africa overall (black).

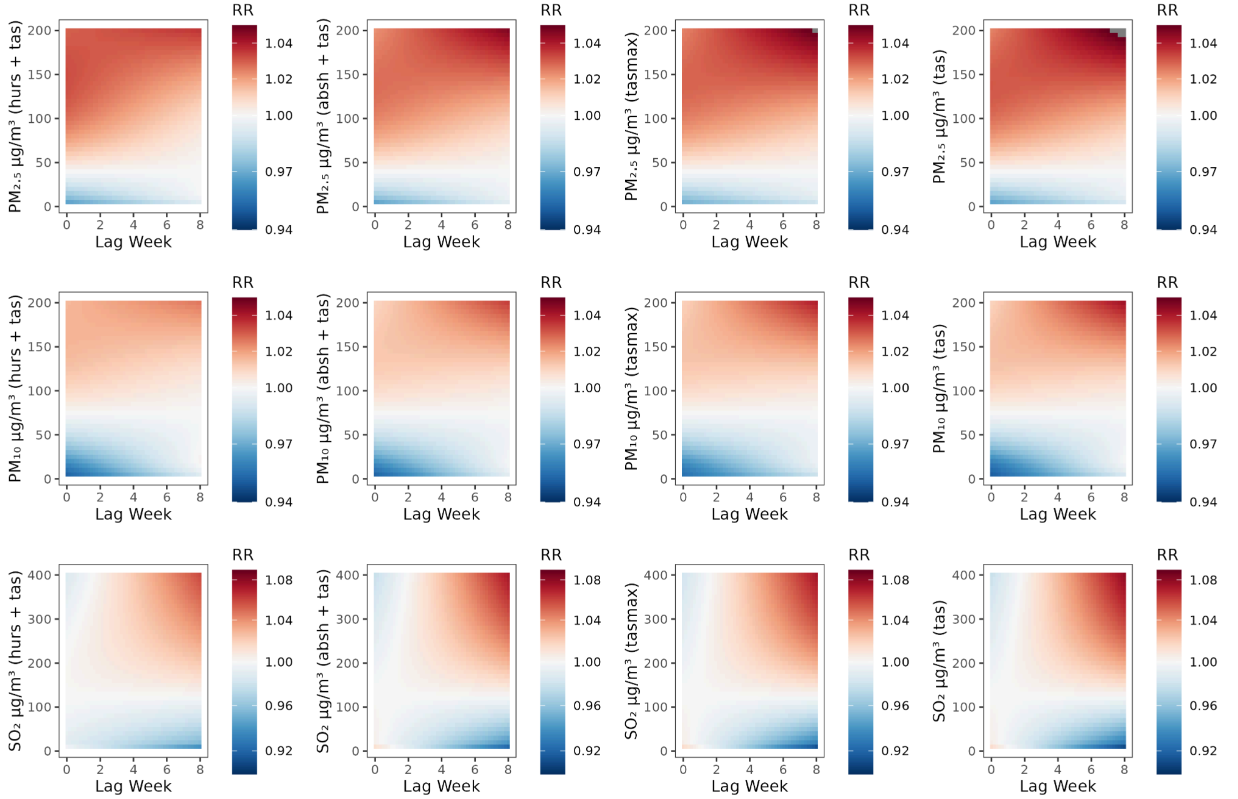


**Figure S12. Median relative risk (RR) surfaces from the air pollution models** where risk >1 is red and <1 is blue for $PM_{2.5}$ (top row), $PM_{10}$ (middle row), and $SO_2$ (bottom row) including adjustment for different formulations of meteorological confounders from left to right for (i) relative humidity and temperature, (ii) absolute humidity and temperature, (iii) maximum temperature, and (iv) mean temperature. The humidity variables are included as non-linear at lag week 3 and the temperature variables are linear at lag week 0. The main air pollution model includes an adjustment for absolute humidity and mean temperature.

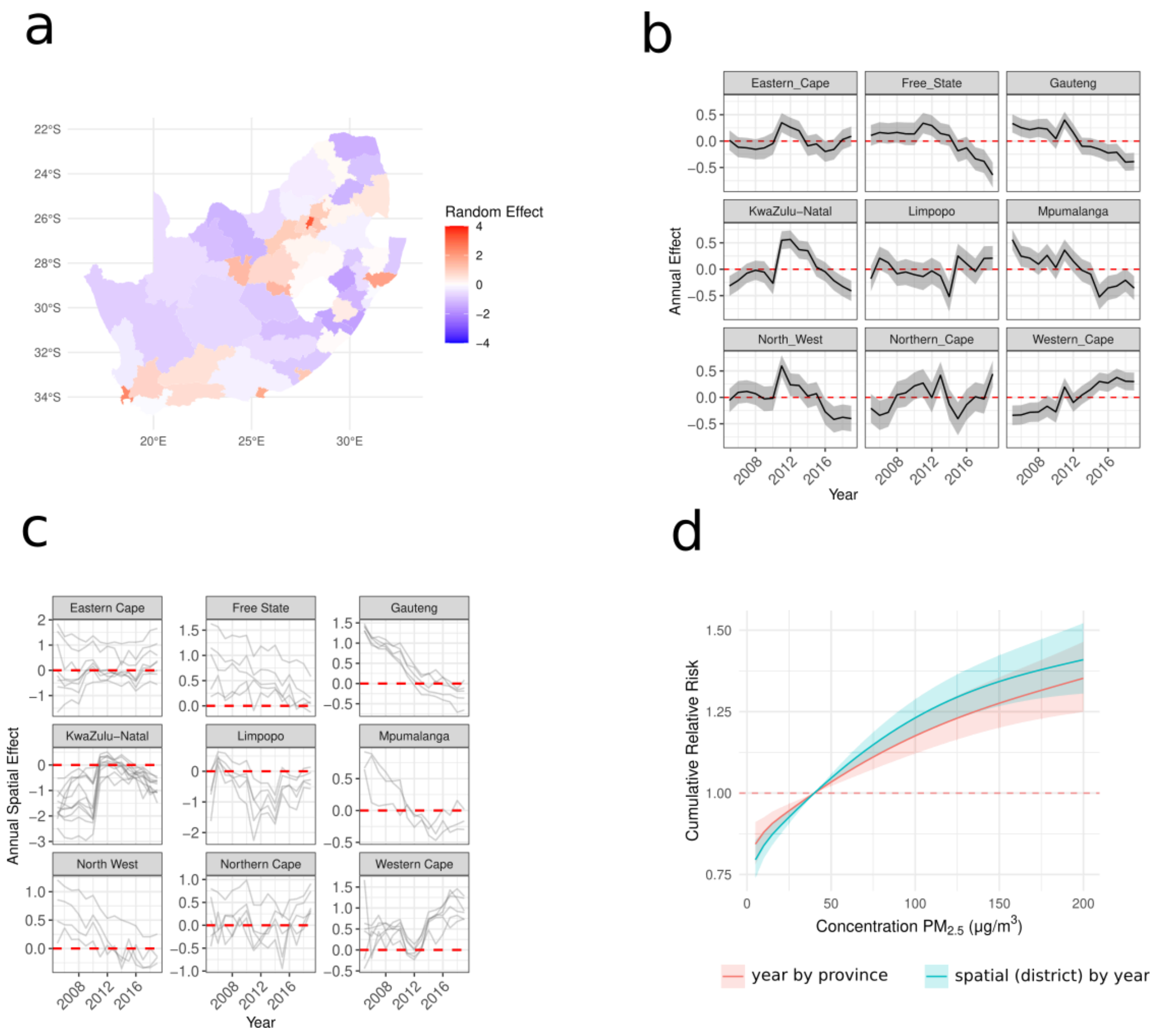


**Figure S13. Model sensitivity with time varying spatial effects.** (a) static spatial random effect from equation 1. (b) interannual effect replicated by province from equation 1. (c) spatial effect replicated by year for each district from equation 2. (d) Cumulative exposure-response curve across 8-weeks with the relative risk (y-axis) across weekly μg/m$^3$ of $PM_{2.5}$ (x-axis) comparing the air pollutant model from the main text (pink) ($\log(\rho_{s,t}) = \alpha + \beta_{pd}pd_{s,a(t)} + f(pcv_{periods}) + \beta_{temp}temp_{s,t} + f(hum_{s,t\text{-}3}) + CB_{aq}(aq_{s,t-\ell}) + \delta_{m(t)} + \gamma_{pr(s),a(t)} + u_s + v_s$) to a model which varies the spatial variation by year (blue) ($\log(\rho_{s,t}) = \alpha + \beta_{pd}pd_{s,a(t)} + f(pcv_{periods}) + \beta_{temp}temp_{s,t} + f(hum_{s,t\text{-}3}) + CB_{aq}(aq_{s,t-\ell}) + \delta_{m(t)} + u_{s,a(t)} + v_{s,a(t)}$).

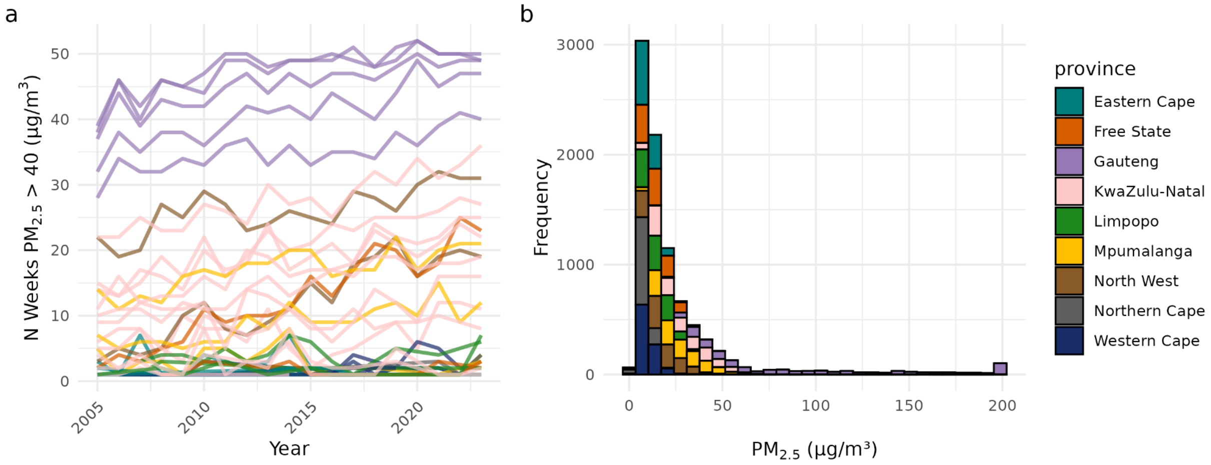


**Figure S14 (Extended Data Figure 6). Particulate matter <2.5 µm distributions** (a) weeks with >40 µg/m$^3$ (the NAAQS threshold for $PM_{2.5}$) per year and district colored by province across the time period. The grouping is by district whereby in Western Cape (dark blue) and Northern Cape (grey) have no weeks exceeding the threshold. Eastern Cape (blue) and Limpopo (green) only have districts with <10 weeks (blue), Free State includes Fezile Dabi (orange), Gauteng includes the City of Johannesburg, City of Tschwane, Ekurhuleni, Sedibeng, and West Rand (purple), KwaZulu-Natal includes Amajuba, eThekwini, Harry Gwala, iLembe, Ugu, uMgungundlovu, uMzinyathi, and uThukela (pink), Mpumalanga includes Gert Sibande and Nkangala with >10 weeks exceeding threshold (yellow), North West includes Bojanala Platinum and Dr Kenneth Kaunda with > 10 weeks (brown) (b) histogram of particulate matter distribution across provinces. Both colored by province.

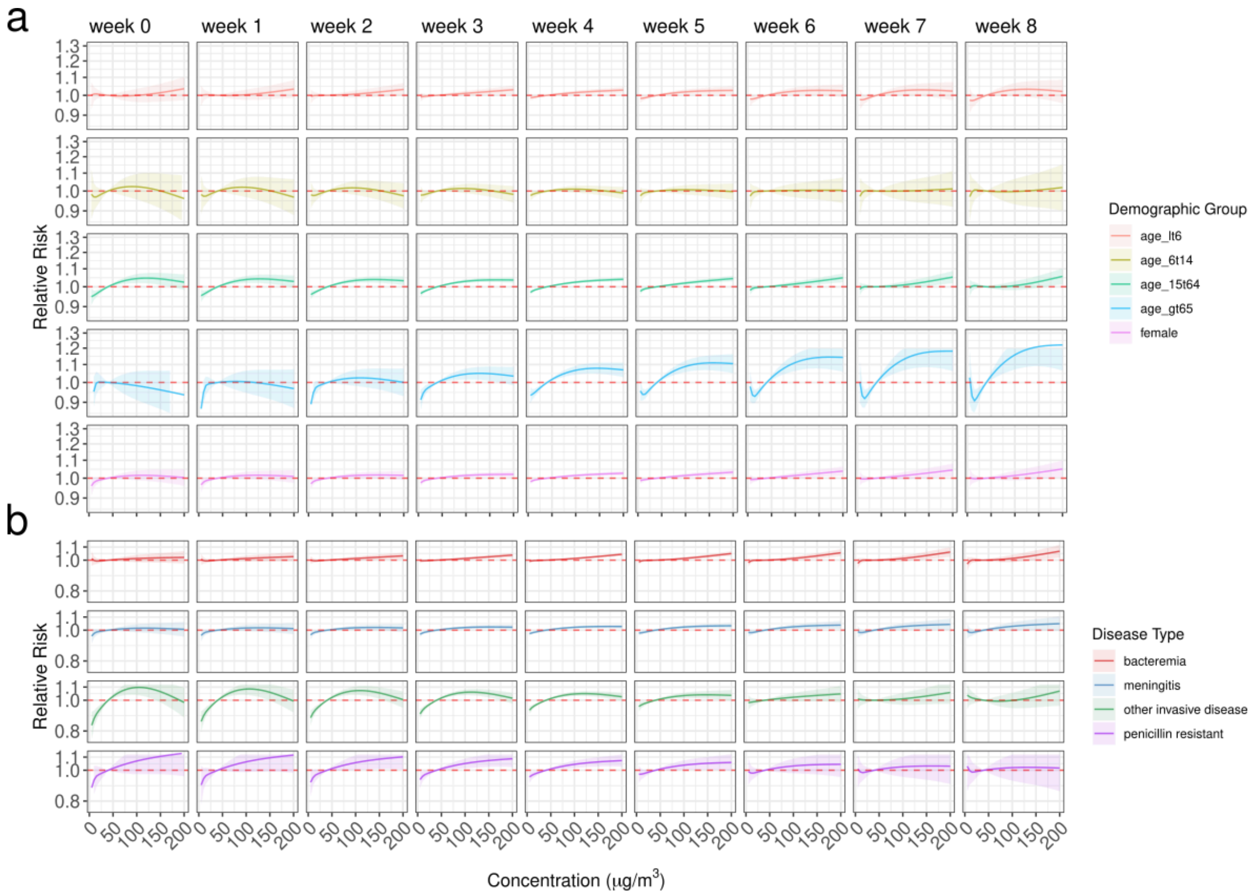


**Figure S15. Exposure-response curves across different weekly exposure to PM$_{2.5}$ (concentration μg/m$^3$) including different stratifications** of invasive disease including (a) demographic factors such as age <6 (red), age 15-64 (green), age >64 (blue), and females alone (purple) and for (b) different disease types including bacteremia (red), meningitis (blue), and other invasive disease types (dark green) all across 8 weeks of lags at the weekly district level.

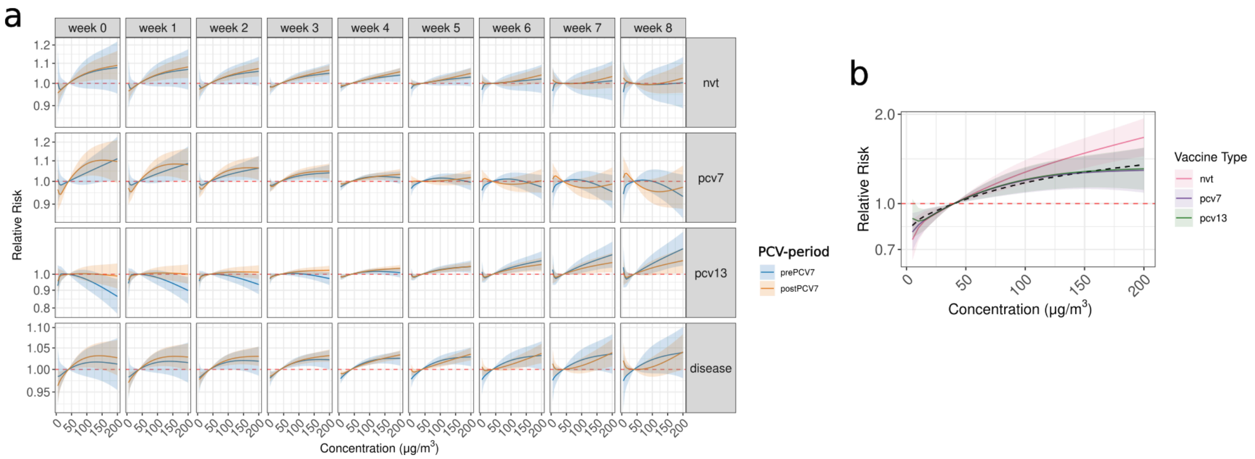


**Figure S16. Exposure-response curves across different weekly exposure to $PM_{2.5}$ (concentration μg/$m^3$)** (a) including different stratifications of invasive disease including that which is caused by NVTs (top), PCV7 (second-down), PCV13 (third-down), or across all disease cases (bottom). These are further stratified by the pre-PCV7 period (2005-2009) (blue) and the post-PCV7 period (2009-2019) (orange) in a model run at the weekly-district level (b) cumulative effect across the entire period for each vaccine group.

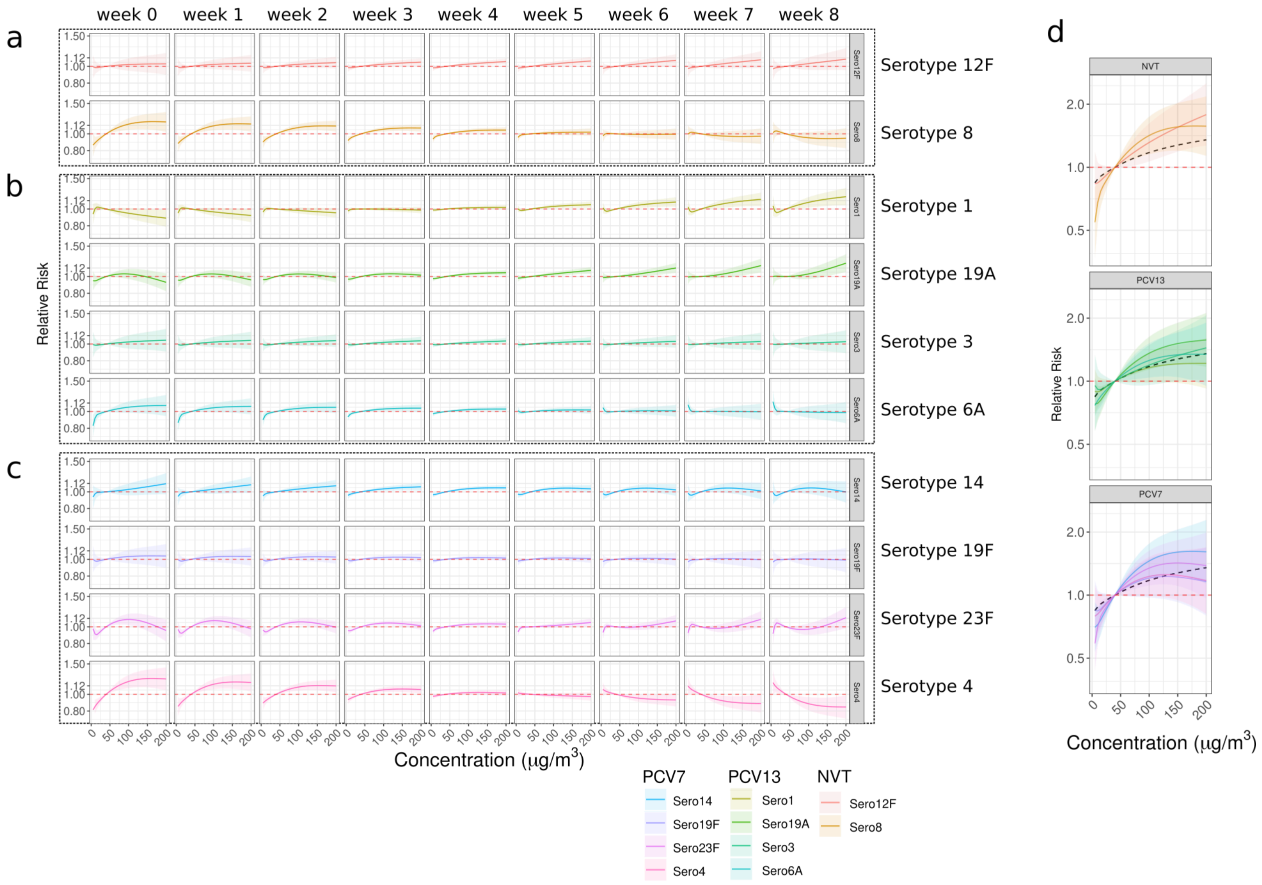


**Figure S17 (Extended Data Figure 7). Exposure-response curves across different weekly exposure to $PM_{2.5}$ (concentration μg/m$^3$) including serotype stratifications** of invasive disease including (a) NVTs serotype 12F and serotype 8, (b) PCV13 serotype 1, serotype 19A, serotype 3, serotype 6A, and (c) PCV7 serotype 14, serotype 19F, serotypes 23F, and serotype 4. (d) includes the cumulative 8-week relative risk for each of these serotype models grouped by vaccine type. The legend is shared across all figures.

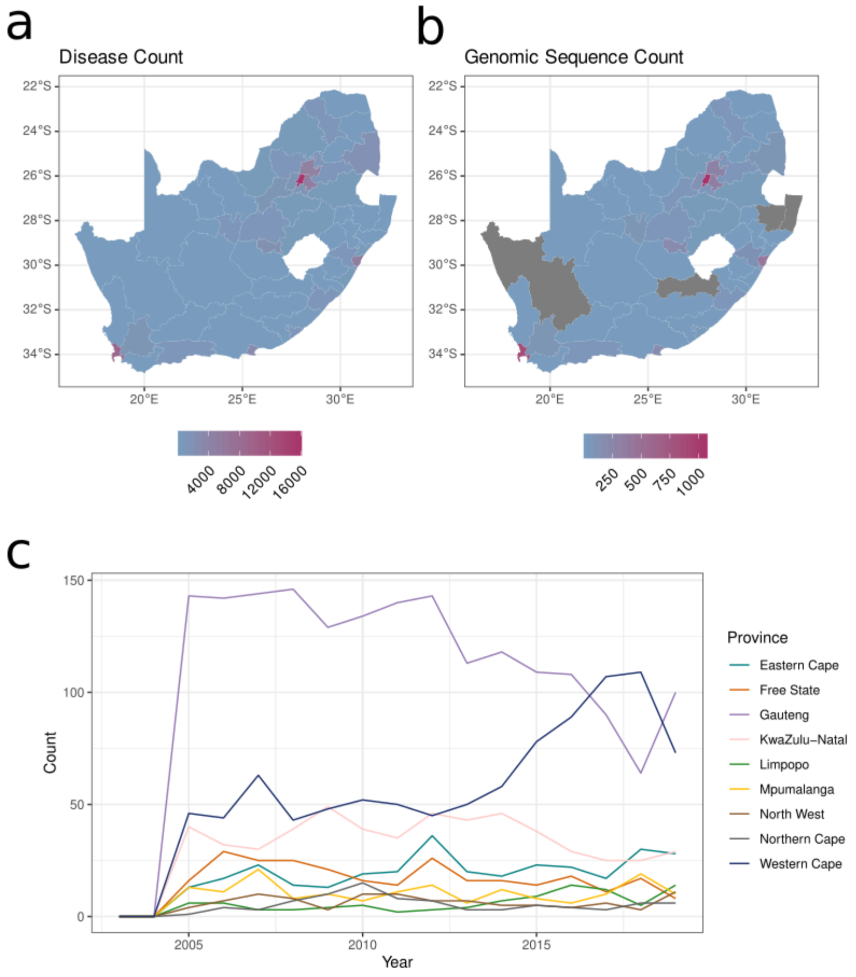


**Figure S18. Summary of GPSC sequences as compared to case count and by province** (a) Number of disease cases from each district and (b) the number of genomic sequences from each district. There were no sequences from 4 districts including: Namakwa, Joe Gqabi, uMkhanyakude, Zululand. (c) The number of pneumococcal samples sequenced from each province over time.

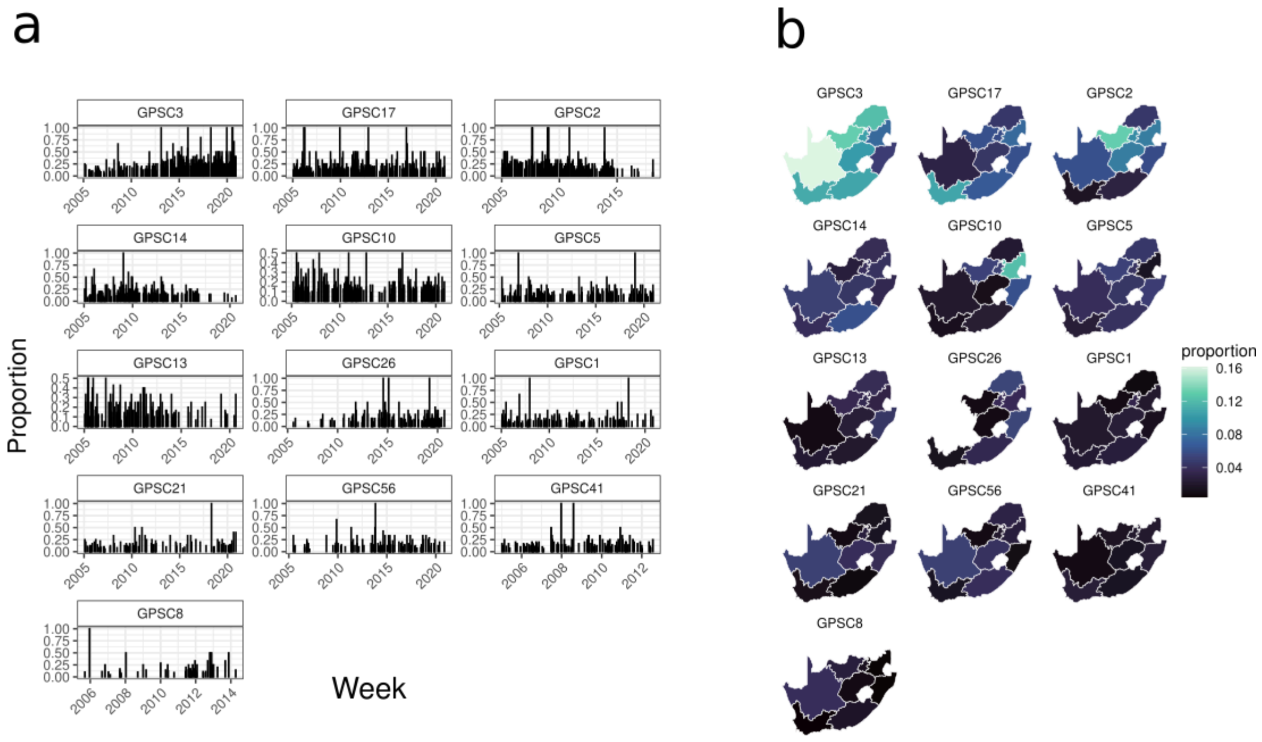


**Figure S19. GPSC proportions** (a) weekly proportion of each GPSC per the total number sequenced in each week across the study period (2005-2023). (b) proportion of each GPSC in each province per the total number sequenced in that province across the study period. GPSCs included are those with N≥100 as well as GPSC8 due to its epidemiological importance in South Africa.

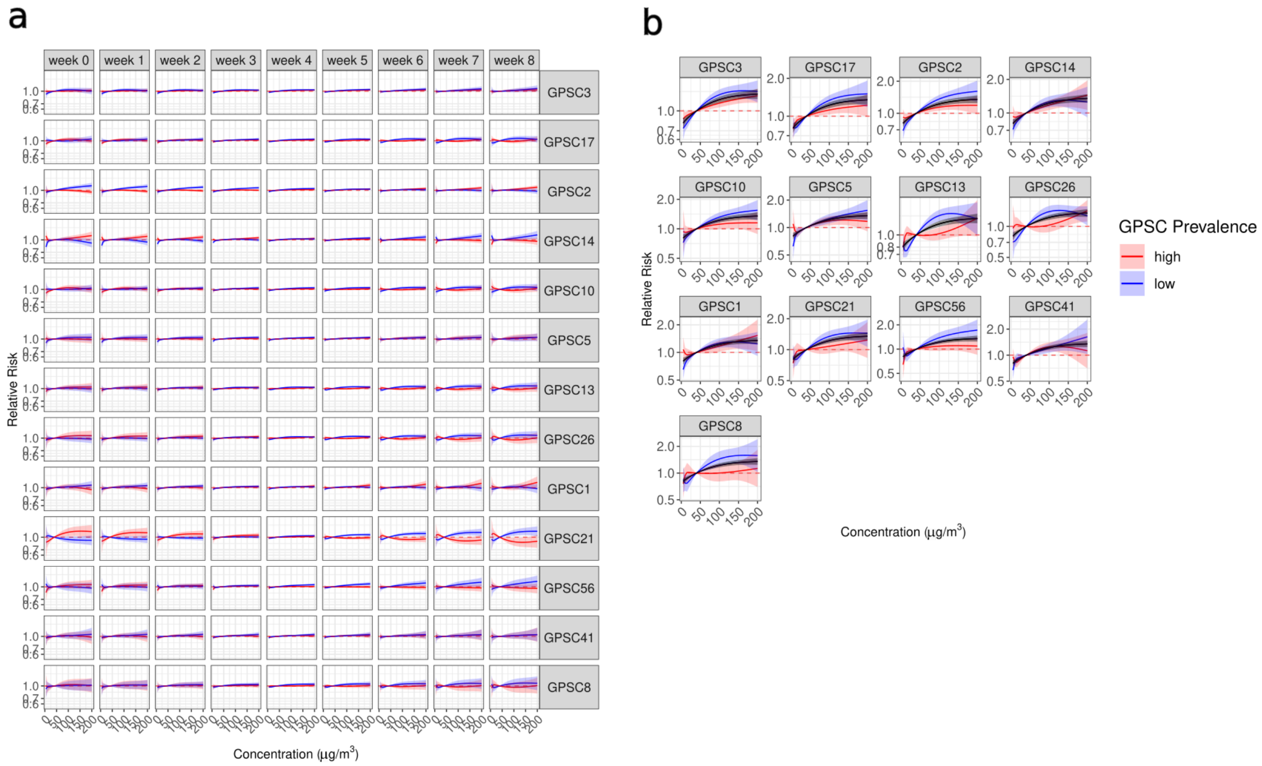


**Figure S20. Exposure-response curves for $PM_{2.5}$ and pneumococcal disease interacted with the proportion of each GPSC per week per province**. (a) 8-week lag time exposure-response curves and (b) includes the cumulative effect. The estimates are at high (red) and low (blue) GPSC prevalence. In b the black line indicates the model with no interaction.

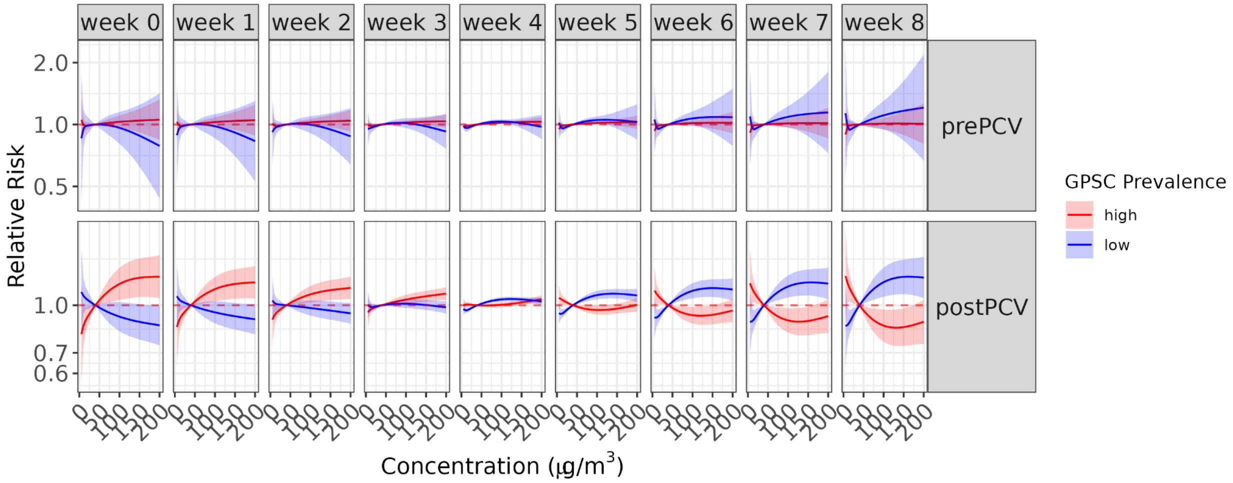


**Figure S21. GPSC21-exposure-response curves for the relative risk of IPD given the concentration of $PM_{2.5}$ and interacted with the weekly proportion of GPSC21** in province level weekly models across an 8-week lag for (top) pre-PCV period 2005-2008 and (bottom) post-PCV period 2009-2019.

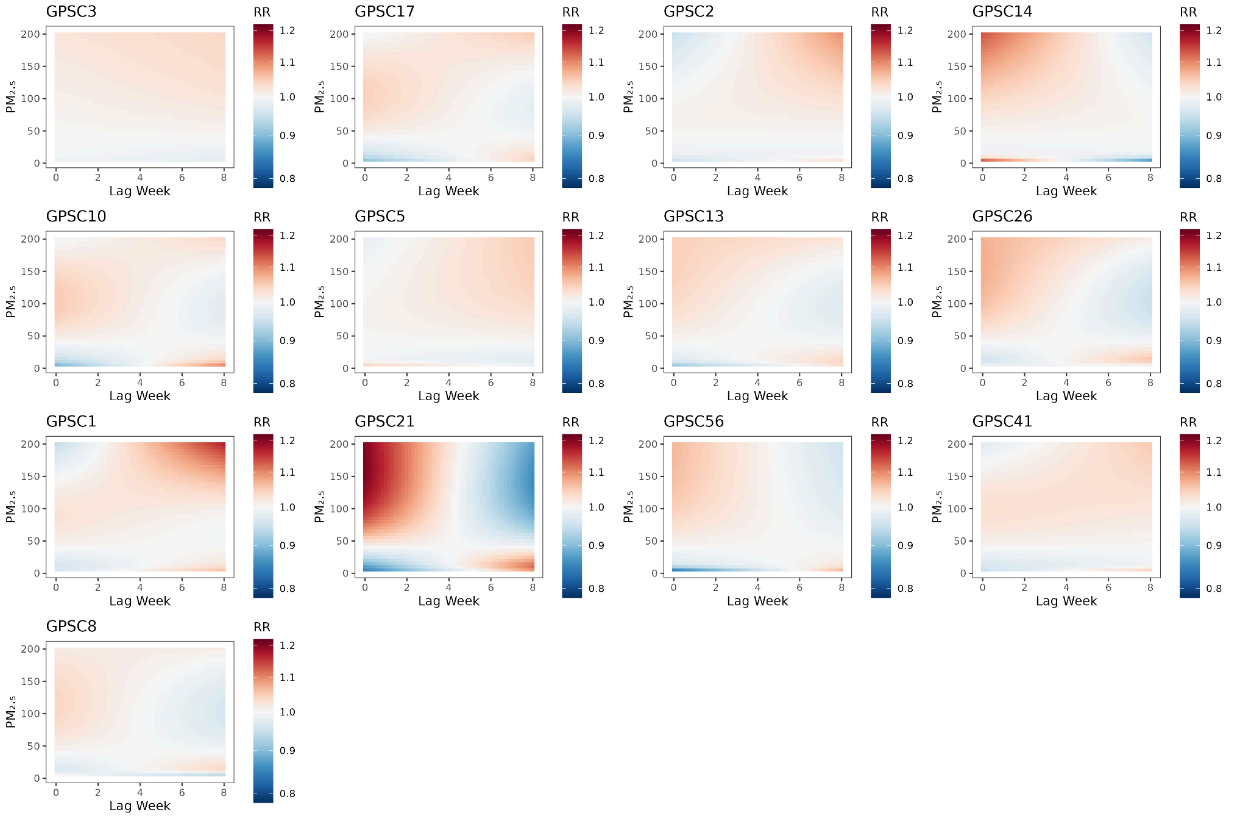


**Figure S22 (Extended Data Figure 8). Median relative risk (RR) surfaces from the air pollution models** where risk >1 is red and <1 is blue with consistent limits across RR's, for different concentrations of $PM_{2.5}$ (y-axis) and 8-weeks of lag (x-axis) when there is high prevalence of each of the GPSCs in order of total count across the time period.

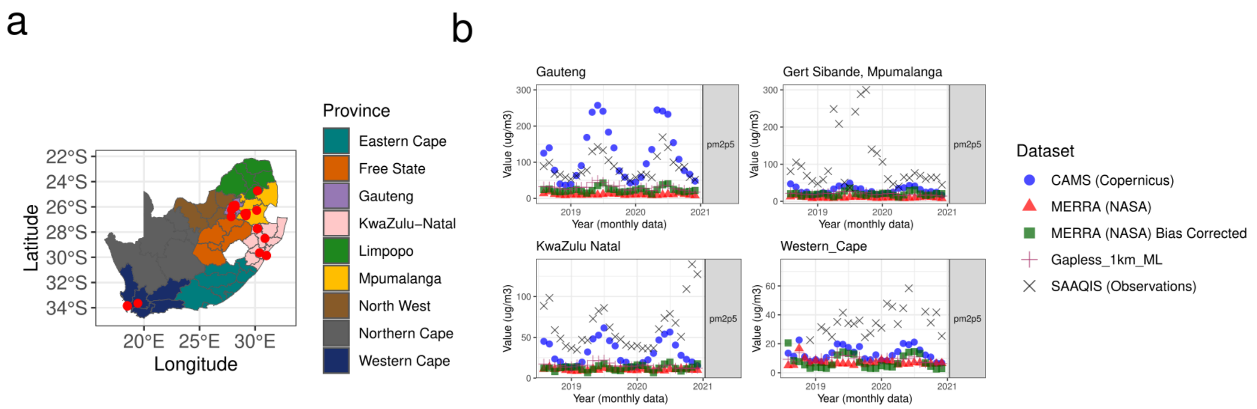


**Figure S23 (Extended Data Figure 9). Reanalysis data compared to observations in South Africa.** (a) Map of air quality monitoring stations included from SAAQIS. Colored by province with lines distinguishing districts. Locations of observation stations are indicated in red dots. (b) The concentration of weekly $PM_{2.5}$ in $\mu g/m^3$ from Gauteng across reanalysis products from Copernicus (CAMS), MERRA-2 from NASA, MERRA-2 with a bias adjustment and a machine learning gapless product at 1km grid squares.
